# Longitudinal Plasma Proteomics Distinguishes Primary Immune Tolerance from Chronic Graft-versus-Host Disease After Pediatric allo-HSCT

**DOI:** 10.64898/2026.10.26.26365891

**Authors:** Daehong Kim, Yaxin Jing, Elli Äärimaa, Erkka Järvinen, Heljä Lång, Adam Alexandersson, Samppa Ryhänen, Minna Koskenvuo, Markku Varjosalo, Eliisa Kekäläinen

## Abstract

Chronic graft-versus-host disease (cGvHD) remains a major cause of morbidity after pediatric allogeneic hematopoietic stem cell transplantation (HSCT), but the early plasma molecular features associated with primary immune tolerance (PIT) versus cGvHD are incompletely understood. We performed longitudinal LC-MS plasma proteomics in 32 pediatric allo-HSCT recipients sampled at Months 1, 3, 6, 9, and 12 after transplantation, quantifying 4,702 proteins. Across timepoint-specific differential-abundance analyses, Month 3 showed the strongest observed PIT-cGvHD proteomic separation and was selected for deeper exploratory characterization. At Month 3, eight proteins were higher in PIT at BH-FDR <0.05, including the 19S proteasome ATPases PSMC1, PSMC3 and PSMC5, the chaperonin subunit CCT3, the ubiquitin-conjugating enzyme UBE2O, and APEH, RAN and NME1, pointing to differences in protein degradation and folding pathways. Time-resolved pathway analysis showed PIT-associated immune, immunoglobulin, and proteostasis programs at distinct post-transplant intervals, whereas Month-3 cGvHD-associated profiles were enriched for platelet activation, cell adhesion, transmembrane transport, and metabolic processes. Outcome-blinded clustering of the 500 most temporally variable proteins resolved three descriptive trajectory patterns: a transient Month-3 peak, a Month-3 nadir with late recovery, and an early-high/late-decline pattern. The distribution of Month-3 PIT–cGvHD protein effects differed across outcome-blinded trajectory clusters, providing an internally consistent temporal framework for the Month-3 signal. These exploratory findings nominate Month 3 as a potentially informative interval of molecular divergence after pediatric allo-HSCT and identify pathway-level features of PIT and cGvHD for prospective validation.

## Introduction

Allogeneic hematopoietic stem cell transplantation (allo-HSCT) is a potentially curative treatment for a range of pediatric malignant and non-malignant disorders. However, its long-term success is substantially limited by graft-versus-host disease (GvHD), particularly the chronic form (cGvHD), which remains the primary driver of non-relapse morbidity and mortality in long-term survivors.^1^ cGvHD is a multisystem immune-mediated disorder arising from dysregulated donor T- and B-cell responses targeting host tissues, and its clinical presentation is highly heterogeneous, ranging from mild organ involvement to severe, therapy-refractory disease.^2^

In contrast, a subset of patients achieves primary immune tolerance (PIT), a state characterized by full donor chimerism, absence of GvHD, and progressive immune reconstitution (IR) without the need for prolonged immunosuppression.^3,4^ Understanding the molecular basis of this divergence between PIT and cGvHD has important implications for risk stratification, therapeutic targeting, and the design of transplant protocols that promote tolerance.^4^

Immune reconstitution after HSCT is a complex, temporally ordered process involving the sequential recovery of innate and adaptive immune cell compartments, thymic output, and B-cell reconstitution, occurring over months to years post-transplant.^5^ Both the pace and quality of immune reconstitution are substantially influenced by the occurrence of GvHD: cGvHD impairs thymic output, delays naïve T-cell recovery, and perturbs B-cell homeostasis, thereby perpetuating immunodeficiency and susceptibility to infections.^6^ Conversely, early and robust immune reconstitution has been associated with reduced GvHD severity and improved overall survival.^7^

The plasma proteome offers a minimally invasive window into systemic immune and inflammatory states. Longitudinal proteomic profiling by liquid chromatography-mass spectrometry (LC-MS) enables unbiased, high-resolution tracking of molecular changes across the post-transplant course and has been applied to identify biomarkers of transplant-related complications, including acute GvHD.^8,9^ However, longitudinal plasma-proteomic studies specifically designed to characterize PIT and cGvHD over the first year after HSCT remain limited. Many studies emphasize cross-sectional or timepoint-specific comparisons, underscoring the need to integrate such analyses with approaches that describe temporal protein patterns across follow-up.

Here, we present an integrated longitudinal analysis of plasma proteomics in a pediatric allo-HSCT cohort. We first characterized the temporal structure of the proteome using outcome-blinded trajectory clustering, then identified Month 3 as the strongest window of PIT-cGvHD divergence. Finally, we tested whether trajectory-defined protein programs are systematically associated with clinical outcome, providing a non-circular framework for interpreting post-transplant proteomic dynamics.

## Materials and Methods

### Patient cohort and clinical classification

This study included 32 pediatric patients who underwent allo-HSCT from the Children”s Hospital, Helsinki University Hospital, Finland, between 2016 and 2022. Patients were followed prospectively with longitudinal plasma sampling at months 1, 3, 6, 9, and 12 post-transplant. Written informed consent was obtained from the parents or legal guardians of all patients, and from the patients themselves when age-appropriate, and the study was approved by the Ethics Committee of the Helsinki University Hospital (HUS/2306/2016). Patients were classified into three outcome groups based on standardized clinical criteria: (1) Primary Immune Tolerance (PIT): no acute GvHD (grade 0) and no cGvHD (n=11); (2) Secondary Immune Tolerance (SIT): acute GvHD present but no cGvHD (n=7); and (3) cGvHD: cGvHD present regardless of acute GvHD status (n=14). Acute and chronic GvHD were graded according to NIH consensus criteria.^3,10^ Among the 14 patients who developed cGvHD, the median time from transplant to cGvHD onset was 3.8 months (range, 3.3-11.6 months); no patient had a documented cGvHD onset before or at the Month-3 sampling window, confirming that Month-3 proteomic sampling preceded clinical cGvHD diagnosis in all cases. Baseline demographics and clinical characteristics are detailed in **Table 1 and Supplementary Figure S1**. Primary comparisons were between PIT and cGvHD; SIT data are presented in supplementary analyses due to limited sample availability.

**Table 1.**
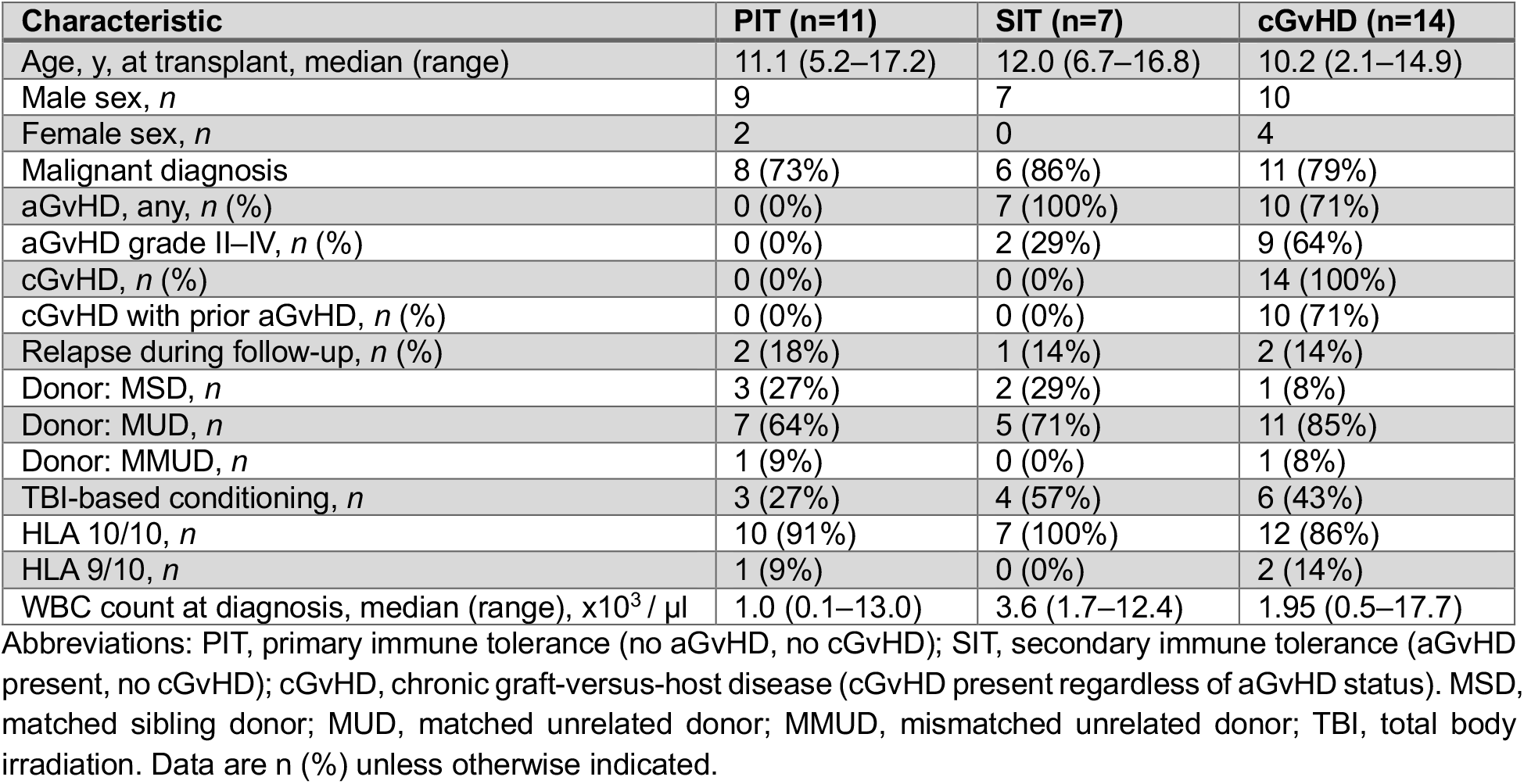
Clinical details of patients.

### Plasma sample collection and LC-MS proteomics

Peripheral blood plasma samples were collected at each timepoint in EDTA-coated tubes. Peripheral blood mononuclear cell isolation was done with gradient centrifugation (Ficoll-Paque PLUS, Cytiva, Buckinghamshire, UK). Plasma was separated by centrifugation at 2000

× g for 15 min and stored at −80°C until analysis. Patients were scheduled for longitudinal sampling at Months 1, 3, 6, 9, and 12 post-transplant; actual sampling completeness varied, with 14 of 32 patients sampled at all five timepoints and 18 patients sampled at 1–4 timepoints (median, 4 timepoints per patient). Month-1 samples were most frequently unavailable. In total,

120 samples were collected and analyzed. Plasma proteins from 20 µl of plasma were enriched with an automated Mag-Net plasma enrichment workflow and the resulting peptides were analyzed with liquid chromatography-mass spectrometry (LC-MS). The fully automated Mag-Net workflow and the LC-MS untargeted proteomics method with Evosep One (Odense, Denmark), employing the 60 samples per day method, connected to Orbitrap Astral (Thermo Fisher Scientific, Waltham, MA, USA) have been previously reported in detail^11^ (**Supplementary Methods**). Method robustness was ensured by processing 2 quality control (QC) samples per plate (**Supplementary Figure S2**), consisting of pooled commercial reference plasma (Research Donors Ltd, London, UK) processed alongside patient samples on the same plates.

### Clinical flow cytometry

Blood immune cell phenotyping was performed on whole blood in the HUS Diagnostic Center clinical laboratory using its validated extended immunophenotyping panel and published gating strategy.^12^ T-cell subsets (CD3, CD4, CD8, CD45RA, CCR7, CD31, TCRαβ, TCRγδ) included naïve (CD45RA^+^CCR7^+^) and recent thymic emigrant (CD45RA^+^CD31^+^) CD4^+^ T cells; B-cell subsets (naïve, memory, switched and non-switched memory, transitional, plasmablast); and NK cells (CD56^+^CD3−) were also defined. Samples were acquired on FACSLyric cytometers and absolute counts were calculated using BD Trucount tubes.

### Trajectory clustering and representative proteins per cluster

For each protein, a cohort mean log2-transformed intensity was calculated at each of the five post-transplant timepoints (Months 1, 3, 6, 9, and 12), averaging across all patients with an available measurement at that timepoint, irrespective of outcome group. Proteins were retained only if a non-missing cohort-mean value was obtained at all five timepoints; no imputation was required, as timepoint-level averaging inherently accommodated per-patient missingness. From the retained proteins, each protein”s five-timepoint cohort-mean trajectory was first z-score standardized (mean 0, SD 1 across the five months), and 500 proteins were selected for downstream clustering based on this standardized representation. The 500 selected proteins were then subjected to k-means clustering (k=3, nstart=100) without using outcome group labels. Cluster-level mean trajectory was computed as the mean z-score per cluster per month. To illustrate the proteins underlying each trajectory pattern, we computed a cluster-specific shape score for each protein quantifying conformity to the cluster-typical profile: for the Month-3-peak cluster, the Month-3 z-score minus the mean z-score of the remaining timepoints; for the Month-3-nadir cluster, the mean of Months 6–12 minus the Month-3 z-score; for the early-high/late-decline cluster, the Month-1 minus Month-12 z-score. The top 12 proteins per cluster by shape score were visualized as a z-score heatmap.

### Differential expression and enrichment analyses

Differential abundance between PIT and cGvHD was assessed independently at each timepoint using limma, with empirical-Bayes moderated t-statistics from an unpaired two-group model (PIT vs. cGvHD); topTable-derived p-values are two-sided. Benjamini–Hochberg FDR correction was applied within each timepoint, and proteins with FDR < 0.05 were considered significant. Proteins were tested if detected in ≥70% of samples at that timepoint; this yielded 1,564, 1,554, 1,678, 1,806, and 1,955 proteins tested at Months 1, 3, 6, 9, and 12, respectively (at Month 3, n_PIT=9, n_cGvHD=13 with available paired data). Missing values among tested proteins were median-imputed per protein before model fitting; for each of the eight FDR-significant Month-3 proteins, at most 2 of 22 samples were imputed (none in PIT). Complete Month-3 results for all tested proteins are provided in **Supplementary Table S2**. To test whether protein trajectories diverged by outcome group across the full follow-up period rather than at isolated timepoints, we additionally fit generalized estimating equation (GEE) models (exchangeable correlation structure, patient-clustered robust standard errors) with a group×time interaction term for each protein with ≥60% completeness across the five timepoints (1,999 of 4,702 proteins). Unlike the per-timepoint limma analyses above, which required ≥70% detection within a single timepoint, this criterion required ≥60% completeness pooled across all five timepoints combined. Benjamini–Hochberg FDR correction was applied across all 1,999 tested proteins. Prior acute GvHD status was not included as a covariate because it is confounded with outcome group (no PIT patients had acute GvHD; 71% of cGvHD patients did). Month 3 was identified as the timepoint of strongest PIT–cGvHD separation in this screen (**Fig. 2A**) and was then characterized in depth (module scores, leave-one-patient-out (LOPO) cross-validation, trajectory-cluster association); pathway enrichment was also examined at Month 1 and 6. Because Month 3 was the maximum of five timepoints, its effect sizes may be modestly inflated.

Gene Ontology Biological Process over-representation analysis (ORA) and gene set enrichment analysis (GSEA) were performed using clusterProfiler (v.4.12.6).^13^ For ORA, the background gene universe was restricted to the set of proteins tested in the corresponding timepoint-specific differential-abundance analysis (rather than the default whole-genome background), ensuring that enrichment was assessed relative to the proteins actually detectable on this platform. For GSEA, the full ranked list of tested proteins at each timepoint (ranked by moderated t-statistic) was used, such that the same timepoint-specific protein universe was implicitly represented.

### Cluster-level association with Month-3 outcome effects

To test whether trajectory-defined programs are associated with clinical outcome without circularity, we compared the distribution of Month-3 log2 fold changes (cGvHD − PIT) across the three trajectory clusters using a linear model (ANOVA), restricted to proteins present in both the trajectory set and the Month-3 differential analysis (n=178). Cluster labels were derived independently of outcome data.

### Statistical and bioinformatic analyses

All analyses were performed in R (v.4.6.1) using the tidyverse (v.2.0.0) for data manipulation, limma (v.3.68.4)^14^ for differential-abundance testing, clusterProfiler (v.4.20.0)^13^ for pathway enrichment, geepack (v.1.3.13)^15^ for generalized estimating equation models, and lme4 (v.2.0.6) for linear mixed-effects models. Figures were generated with ggplot2 (v.4.0.3) and ggrepel (v.0.9.8). Multiple testing across proteins was controlled with the Benjamini–Hochberg false discovery rate (FDR); the thresholds used are given in each Methods subsection. Two-sided tests were used throughout unless stated otherwise.

## Results

### Cohort characteristics and proteomic profiling

The study cohort comprised 32 pediatric allo-HSCT recipients (PIT n=11, SIT n=7, cGvHD 14), sampled longitudinally at Month 1, 3, 6, 9, and 12 after transplantation (**Fig. 1A; Table 1**). Median age at transplant was 10.8 years (range, 2.1-17.2). Donor source, conditioning regimen, and HLA-matching status are shown in **Table 1**. Acute GvHD occurred in 17 patients (53%); 10 of them progressed to cGvHD, and 4 further patients developed cGvHD without preceding acute GvHD (total cGvHD n=14, 44%). White blood cell counts at diagnosis were 1.0, 1.95, and 3.6 x10^3^/µL in PIT, cGvHD and SIT, respectively (**Table 1**), mainly reflecting underlying diagnosis (non-malignant 4.4 vs. malignant 1.7 x10^3^/µL). LC-MS profiling quantified 4,702 plasma proteins across 120 samples from 32 patients. The median number of protein identifications per sample was 2,336 (range, 1,157–3,915), corresponding to a median per-sample detection rate of 49.7% of the total quantified proteome. Identification depth was consistent across timepoints (median, 2,240–2,533 proteins per sample; **Supplementary Figure S2**).

**Figure 1.**
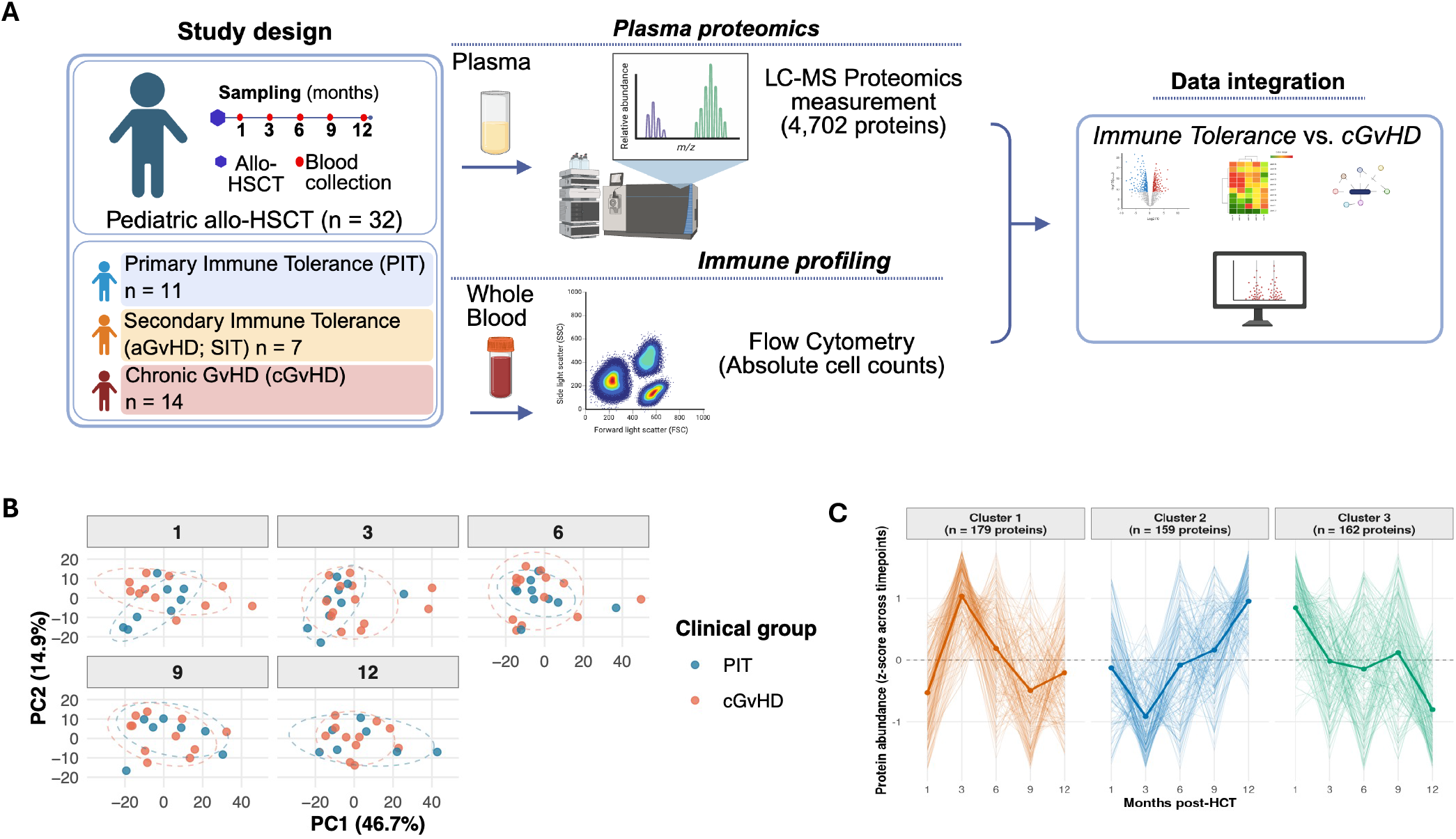
Study design and unsupervised structure of the longitudinal plasma proteome after pediatric allo-HSCT. **(A)** Study design. Thirty-two pediatric allo-HSCT recipients were classified as primary immune tolerance (PIT, n=11), secondary immune tolerance (SIT, n=7), or chronic GVHD (cGVHD, n=14) and sampled at Months 1, 3, 6, 9, and 12 post-transplant for LC-MS plasma proteomics and flow cytometry. **(B)** Global plasma-proteome structure across post-HSCT timepoints. PCA of the top most variable proteins, faceted by timepoint. PC1 and PC2 explain 46.7% and 14.9% of variance. Ellipses: 95% confidence regions per group. **(C)** Outcome-blinded trajectory clustering of the 500 most variable proteins (within-protein z-scores across timepoints, k=3). Thin lines: individual proteins; thick lines: cluster means.

**Figure 2.**
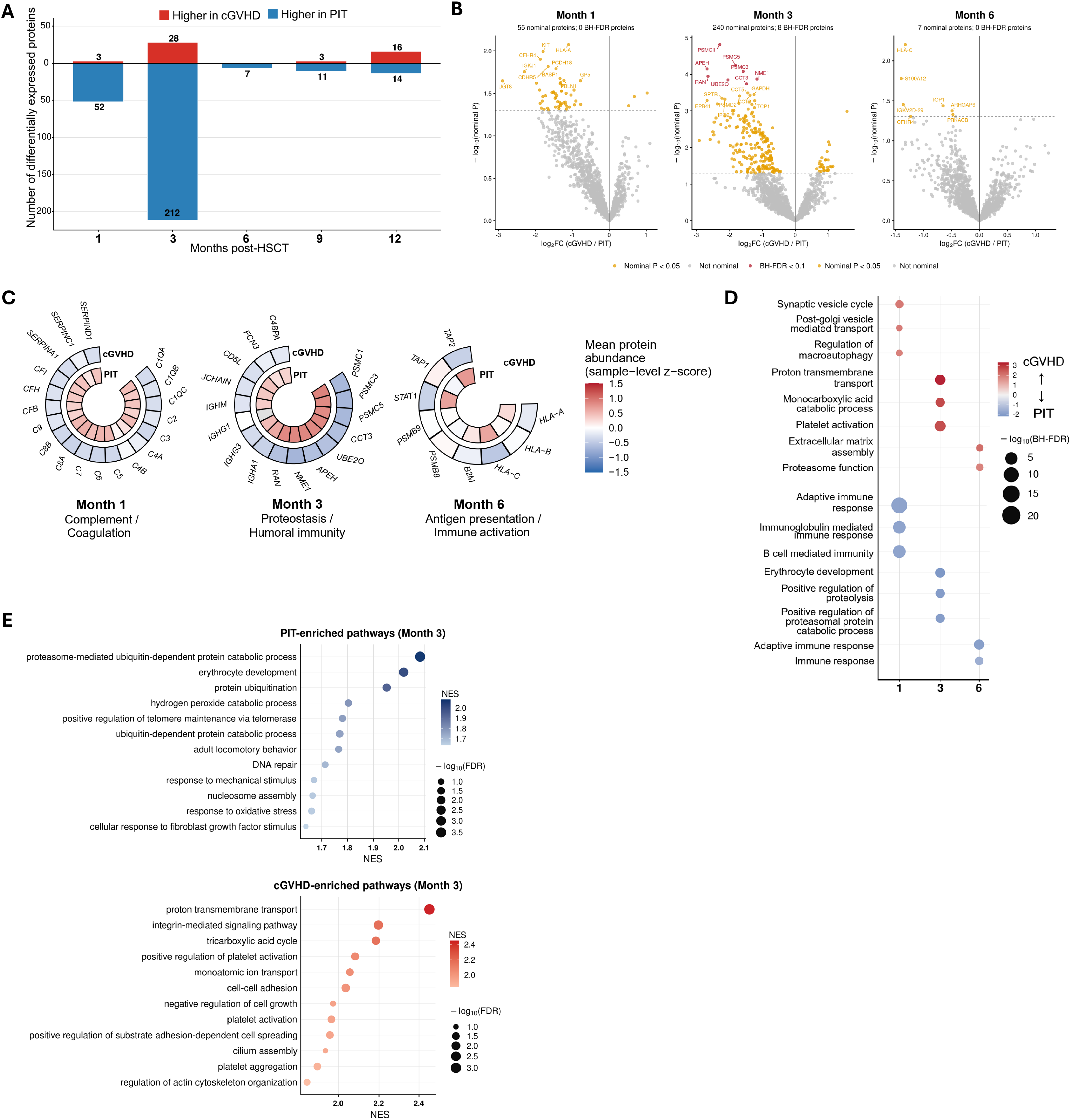
PIT and cGVHD diverge most strongly at Month 3 through a coordinated proteostasis program. Number of differentially abundant proteins between PIT and cGVHD at each post-HSCT timepoint, showing Month 3 as the peak window of proteomic separation. **(B)** Volcano plot of Month-3 plasma protein abundance differences between cGvHD and PIT (limma moderated t-test; 1,554 proteins tested; positive log2 fold change = higher in cGvHD). Eight PIT-higher proteins met BH-adjusted FDR < 0.05 (red): PSMC1, PSMC5, APEH, PSMC3, RAN, NME1, UBE2O and CCT3. Orange points indicate proteins with nominal P < 0.05 (n = 240) that did not reach FDR < 0.05; grey points were not significant. Complete results for Month 3, including the number of observed and imputed values per protein, are provided in **Supplementary Table S2. (C)** Circular heatmaps of curated protein sets: complement/coagulation (Month 1), proteostasis/humoral immunity (Month 3) and antigen presentation/immune activation (Month 6). The Month-3 set comprises the eight FDR < 0.05 proteins from panel B plus IGHA1 and IGHG3. The proteostasis/humoral-immunity module was specifically elevated in PIT at Month 3, converging with the individual-protein (Panel B) and unbiased pathway-enrichment (Panel D) results below. **(D)** Unbiased, timepoint-specific GO-BP gene set enrichment analysis (all tested terms, no pre-selection), independently confirming proteostasis- and ubiquitin-pathway enrichment among PIT-associated proteins at Month 3. **(E)** Month-3 GO-BP gene-set enrichment, shown separately for pathways enriched in PIT (top, blue) and in cGVHD (bottom, red).

### Global proteomic landscape is dominated by inter-individual and temporal variations

To characterize the overall proteomic structure, we performed principal component analysis (PCA) on the 577 proteins quantified in all 100 PIT and cGvHD samples (**Fig. 1B**). PIT and cGvHD samples overlapped substantially at every timepoint, and neither outcome group, timepoint nor patient identity explained appreciable variance in PC1 or PC2 (adjusted R^2^ ≤ 0.04; PC1 and PC2 explained 46.7% and 14.9% of total variance). PC1 tracked the number of proteins identified per sample (Spearman ρ = −0.92), indicating a depth-related axis. In a PCA of the 200 most variable proteins PC1 55.0%, PC2 17.2%; **Supplementary Fig. S3A, B**), same-patient samples were not significantly closer than different-patient samples in PC1–PC2 space (median distance 12.5 vs. 12.9; Wilcoxon P = 0.41; **Supplementary Fig. S3C**). The absence of outcome-related structure in the leading components supported targeted longitudinal analyses rather than relying on unsupervised separation alone.

To define temporal protein patterns independently of clinical outcome, proteins were z-score standardized across timepoints and the 500 most variable proteins were clustered according to their longitudinal profiles. Outcome-blinded k-means clustering with three prespecified clusters resolved three principal trajectory patterns (**Fig. 1C**): a transient Month-3 peak (Cluster 1, n=179), a Month-3 nadir with progressive late recovery (Cluster 2, n=159), and an early-high, late-decline pattern (Cluster 3, n=162).

We subsequently projected samples from patients with secondary immune tolerance (SIT) onto this proteomic framework. PCA, signature-score, and cluster-trajectory analyses were used descriptively to compare SIT with PIT and cGvHD across follow-up (**Supplementary Fig. S4**). These analyses showed broad overlap among the three outcome groups and were not used to identify additional FDR-significant proteins or to establish a definitive SIT-specific proteomic phenotype, given the exploratory design and limited SIT sample size.

### PIT and cGvHD diverge most strongly at Month 3, with higher abundance of proteasome-, chaperonin- and ubiquitin-related proteins in PIT

We examined when PIT–cGvHD proteomic differences were greatest and which biological programs characterized the divergence. Timepoint-specific differential-abundance analysis showed Month 3 as the strongest window of PIT–cGvHD separation (**Fig. 2A**; volcano plots for all timepoints in **Supplementary Fig. S5**). In a group×time interaction model, 7 of 1,999 proteins with sufficient longitudinal completeness showed a significant interaction (FDR<0.05; **Supplementary Table S1, Supplementary Fig. S6**), all declining over follow-up in PIT and rising in cGvHD. Because time was modeled linearly, this analysis detects sustained average differences and is complementary to, not confirmatory of, the transient Month 3 signal (**Fig. 1C, 2, and 4**).

At Month 3, 240 of 1,554 tested proteins differed between cGvHD and PIT at nominal P < 0.05, and eight remained significant after Benjamini–Hochberg correction (BH-FDR < 0.05; **Fig. 2B; Supplementary Table S2**). All eight were higher in PIT: PSMC1, PSMC5 and PSMC3 (19S proteasome ATPase subunits), CCT3 (a chaperonin subunit), UBE2O (an E2/E3 ubiquitin ligase), and APEH, RAN and NME1. Abundance was 2.2-to 6.4-fold lower in cGvHD. Further chaperonin and proteasome proteins, including CCT5, CCT2 and PSMD2, ranked just below the threshold (BH-adjusted P = 0.050). No protein met BH-FDR < 0.05 at Months 1, 6, 9 or 12.

To examine whether this signal extended beyond individual proteins, we summarized the abundance of curated protein modules. The Month 1 and Month 6 modules were selected from GO-BP categories, whereas the Month 3 module comprised the eight FDR-significant proteins from **Fig. 2B** together with IGHA1 and IGHG3; these circular heatmaps are therefore descriptive summaries and not independent tests (**Fig. 2C**). The predominant group-level pattern changed over time: complement/coagulation proteins were displayed at Month 1, a proteostasis/humoral-immunity set at Month 3, and antigen-presentation/immune-activation proteins at Month 6. In particular, the Month-3 circular heatmap showed higher relative abundance of multiple proteostasis-associated proteins in PIT, including PSMC1, PSMC3, PSMC5, CCT3, UBE2O, RAN, NME1, and APEH. These descriptive module summaries support temporally structured molecular differences between PIT and cGvHD, rather than a single invariant group signature across follow-up.

As an unbiased complement to the curated modules above, we performed timepoint-specific, unbiased GO-BP gene set enrichment analysis (**Fig. 2D**). At Month 1, PIT-associated enrichment was dominated by adaptive immune response (NES = −1.88, BH-FDR = 3.68 x 10^-23^), immunoglobulin-mediated immune response (NES = −1.86, BH-FDR = 1.03 x 10^-11^), and complement activation^16^ (NES = −1.65, BH-FDR = 1.96 x 10^-4^). In contrast, the strongest cGvHD-associated Month-1 terms were synaptic vesicle cycle (NES = 2.09, BH-FDR = 0.0083), regulation of macroautophagy (NES = 1.94, BH-FDR = 0.035), and post-Golgi vesicle-mediated transport (NES = 2.04, BH-FDR = 0.037).

At Month 3, the timepoint of strongest divergence, cGvHD-associated enrichment included platelet activation^17^ (NES = 2.79, BH-FDR = 8.11×10^-7^), proton transmembrane transport (NES = 3.30, BH-FDR = 6 × 10^-6^), monocarboxylic-acid catabolic process (NES = 2.85, BH-FDR = 4.2 × 10^-4^), and platelet aggregation (NES = 2.49, BH-FDR = 1.88 × 10^-4^). In contrast, and consistent with both the individual-protein (**Fig. 2B**) and module-level (**Fig. 2C**) results above, the PIT-associated profile showed strong enrichment of proteasome-mediated ubiquitin-dependent protein catabolism (NES = −1.89, BH-FDR = 2.45 x 10^-5^), erythrocyte development (NES = −2.15, BH-FDR = 4.79 x 10^-5^), and ubiquitin-dependent protein catabolism (NES = −1.78, BH-FDR = 5.43 x 10^-5^).

At Month 6, PIT-associated immune signatures remained prominent, including adaptive immune response (NES = −1.97, BH-FDR = 1.15 x 10^-5^) and immune response (NES = −1.59, BH-FDR = 0.0032). Conversely, cGvHD-associated enrichment included proteasome-mediated ubiquitin-dependent protein catabolism (NES = 1.92, BH-FDR = 0.0177) and extracellular-matrix assembly (NES = 2.27, BH-FDR = 0.0258). Together, these three complementary lines of evidence (individual proteins (**Fig. 2B**), pre-specified modules (**Fig. 2C**), and unbiased pathway enrichment (**Fig. 2D**)) converge on the same conclusion: functional separation between PIT and cGvHD is time-dependent, with the most coherent and extensive opposing pathway signals observed specifically at Month 3.

Given this convergent evidence for Month 3 as the clearest window of coordinated divergence, we examined the corresponding pathway architecture in detail. At this timepoint, the PIT-associated ranking was enriched for protein-catabolism and proteostasis-related processes, including proteasome-mediated ubiquitin-dependent protein catabolic process, erythrocyte development, protein ubiquitination, and hydrogen peroxide catabolism (**Fig. 2E**; **Supplementary Table S3**). In contrast, the cGvHD-associated ranking was enriched for proton transmembrane transport, tricarboxylic-acid-cycle and monocarboxylic-acid-catabolic processes, platelet activation and aggregation, cell–cell adhesion, and integrin-related signaling.^17^ Together, these findings indicate that the Month-3 proteomic contrast reflects coordinated differences between PIT-associated protein-quality-control programs and cGvHD-associated transport, metabolic, platelet, and cell-adhesion programs.

### Month 3 PIT proteomic and immune reconstitution

We next asked whether the Month-3 PIT proteomic program related to immune reconstitution. A composite PIT pathway score, built from standardized abundance of proteins in the top PIT-enriched GO biological-process pathways at Month 3, was higher in patients subsequently classified as PIT than in those with cGvHD (**Fig. 3A**). To address the circularity, we used leave-one-patient-out (LOPO) cross-validation, re-deriving the signature without each patient before scoring that patient. The PIT-associated signature remained significantly higher in PIT (Mann-Whitney P=0.043, **Fig 3E**), whereas the same procedure applied to the Month 1 and Month 6 PIT-associated signatures was not significant (Month 1: P = 0.44, n = 19 [PIT n = 8, cGvHD n = 11]; Month 6: P = 0.21, n = 23; **Supplementary Table S4**). No reciprocal cGvHD-associated signature was available for Months 1 or 6, so this comparison is limited to the PIT direction. The pattern suggests that the PIT-associated program is localized to the Month-3 window rather than reflecting a persistent baseline difference. In patients with paired Month-3 proteomic and flow-cytometry data, higher PIT pathway scores showed nominal positive associations with absolute counts of recent thymic emigrants (RTE) and naïve CD4^+^ T cells, and weaker or inconsistent associations with naïve CD8^+^ T cells, naïve B cells, and γδ T cells (**Fig. 3B**). Confidence intervals were wide and analyses unadjusted, so these findings are exploratory. Median trajectories of these subsets over Months 1–12 suggested numerically greater recovery of RTE and naïve CD4^+^ T cells in PIT, whereas the other subsets were inconsistent (**Fig. 3C**). Linear mixed-effects models (group, month and their interaction; patient random intercept; log10(cells/µL^+^1)) pointed in the same direction, but confidence intervals were wide and generally included the null, so there was no definitive evidence of differential longitudinal reconstitution between clinical groups (**Fig. 3D**). To obtain an exploratory comparison in an independent cohort, we analyzed an external pediatric allo-HSCT cohort previously profiled using targeted Olink Inflammation and Immune Response panels.^18^ None of the eight proteins prioritized by LC-MS was measured on these panels, and only nine of the 180 Olink proteins were also tested in the LC-MS Month 3 analysis, so this analysis could not replicate the LC-MS signal. At Month 3, 27 of 180 proteins had nominal P<0.05 (about nine expected by chance), with higher TRANCE, NCR1, CD83, and CD28 in PIT and higher AREG, OPG, IL-18R1, IL8, and IL10 in cGvHD (**Supplementary Fig. S7; Supplementary Table S5**). No protein passed FDR<0.05 (smallest BH-adjusted P=0.074, TRANCE). These results are therefore hypothesis-generating and do not constitute replication, given the small cohort, targeted assay design, and differences protein coverage compared with untargeted LC–MS.

**Figure 3.**
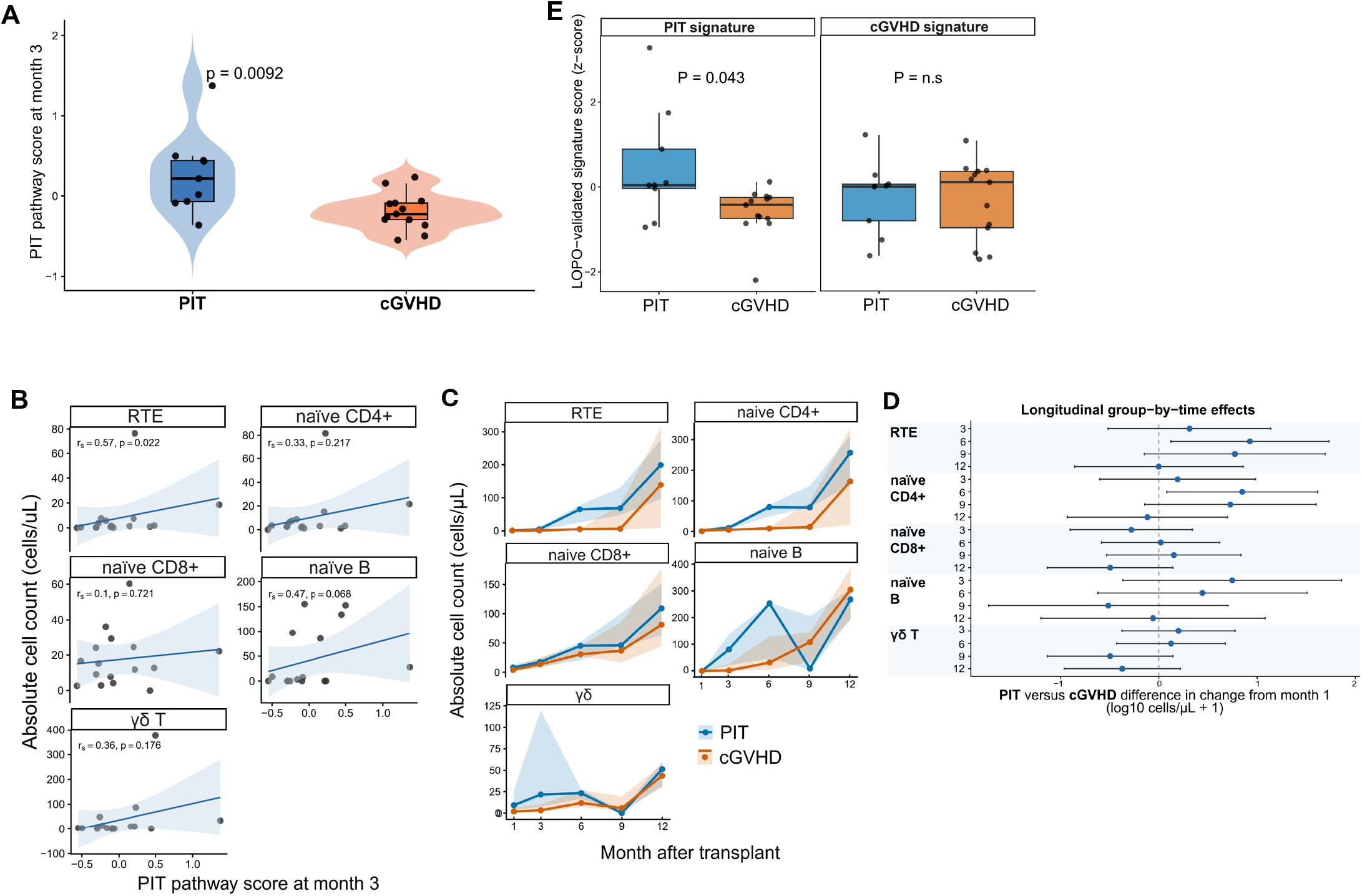
**(A)** Composite PIT pathway score at Month 3, calculated from standardized abundance of proteins mapping to top PIT-enriched GO-BP pathways, compared between patients subsequently classified as PIT versus cGVHD. LOPO cross-validation (**Fig. 3E**), in which the signature was re-derived excluding each scored patient, confirmed this difference (Mann-Whitney P=0.043), indicating the signal is not attributable to circular scoring. **(B)** Association of month-3 PIT pathway score with absolute counts of recent thymic emigrants (RTE), naïve CD4^+^ T cells, naïve CD8^+^ T cells, naïve B cells, and γδ T cells. Each point represents one patient with paired proteomic and flow-cytometry data; lines show linear fits with 95% CIs; annotations show Spearman ρ and unadjusted P values. **(C)** Longitudinal absolute counts of RTE and naïve T- and B-cell subsets at months 1, 3, 6, 9, and 12 post-HCT, stratified by PIT and cGVHD. Lines and shaded bands show median and interquartile range; numbers of evaluable patients at each timepoint are indicated. **(D)** Model-based estimates of differential longitudinal reconstitution between PIT and cGVHD from linear mixed-effects models for each cell subset. Points show estimated differences in change from month 1 (PIT minus cGVHD) at months 3, 6, 9, and 12; horizontal bars indicate 95% CIs. Positive values indicate greater relative recovery in PIT. **(E)** LOPO cross-validation of the Month-3 PIT pathway score shown in Panel A. The signature was re-derived excluding each patient in turn and used to score that patient independently, confirming the PIT-associated signal is not attributable to circular scoring (Mann-Whitney P=0.043).

### Trajectory clustering reveals a coordinated month 3 proteomic program

Because single-protein analysis at Month 3 has limited power, we defined trajectory clusters from temporal patterns across all timepoints, without clinical labels. This yielded three patterns: a transient month-3 peak, a month-3 nadir with later recovery, and an early-high, late-decline pattern (**Fig. 4A**). We next summarized these patterns at the pathway level. Mean PIT and cGvHD proteomic signature scores (z-scores) showed that the PIT signature was higher in patients subsequently classified as PIT, particularly around month 3, whereas the cGvHD signature showed the opposite pattern (**Fig. 4B**). Under the same LOPO validation (**Fig. 3E**), the PIT-associated signature remained nominally higher in PIT (P=0.043), whereas the reciprocal cGvHD-associated signature did not (P=0.42). Within each trajectory cluster, we ranked proteins by a trajectory shape score and selected the top 12 per cluster. These representative proteins recapitulated the cluster-level trajectories and spanned immune, complement, and metabolic functions, suggesting coordinated programs rather than isolated changes (**Fig. 4C**). Cross-referencing these representative proteins against Human Protein Atlas immune cell transcriptomic data showed no statistically significant enrichment for a specific immune cell lineage within any cluster (**Supplementary Fig. S8, Supplementary Table S6**), though several proteins (e.g., CEMIP, CFI, CHRD) are not expressed by circulating immune cells and may represent non-leukocyte-derived plasma components. We then tested whether trajectory cluster membership predicted Month-3 differential abundance between PIT and cGvHD. Cluster membership was associated with Month-3 log fold changes (ANOVA F(2,175) = 14.56, p = 1.4 × 10^−10^) (**Fig. 4D**), with Clusters 1 and 3 higher in PIT and Cluster 2 higher in cGvHD. Because cluster assignment incorporates each protein”s Month-3 shape, this consistency is not independent replication of the Month-3 finding.

**Figure 4.**
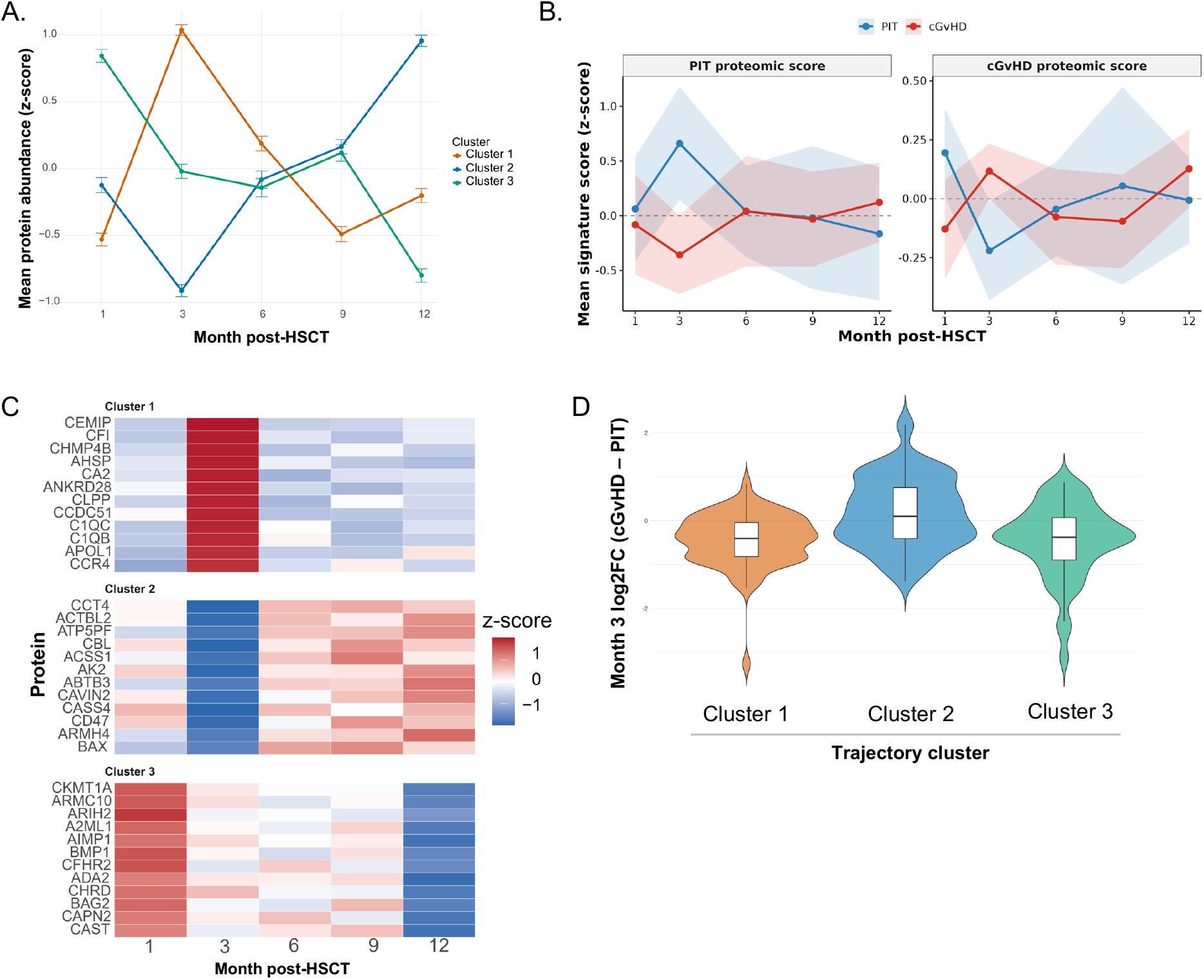
Longitudinal proteomic signatures and trajectory-defined molecular programs. **(A)** Mean protein abundance (z-score) for three outcome-blind trajectory clusters across months 1, 3, 6, 9, and 12 post-HSCT. Points and error bars indicate mean ± SE. Cluster 1 shows a transient month-3 peak, Cluster 2 a month-3 nadir with later recovery, and Cluster 3 an early-high, late-decline pattern. **(B)** Mean PIT and cGVHD proteomic signature scores (z-scores) across Months 1–12, by outcome group. LOPO cross-validation confirmed the PIT-associated signature (P=0.043) but not the cGVHD-associated signature (P=0.42), indicating asymmetric robustness between the two signatures. **(C)** Representative proteins for each trajectory cluster. Heatmap of cohort-mean z-scored abundance across Months 1, 3, 6, 9 and 12 for the 12 proteins with the highest cluster-specific shape score in each of the three outcome-blinded clusters (36 proteins in total). Shape scores measure conformity to the cluster-typical profile: for the Month-3-peak cluster (Cluster 1), the Month-3 z-score minus the mean z-score of the remaining timepoints; for the Month-3-nadir cluster (Cluster 2), the mean of the Month 6–12 z-scores minus the Month-3 z-score; and for the early-high, late-decline cluster (Cluster 3), the Month-1 minus Month-12 z-score. Rows are proteins grouped by cluster and columns are timepoints. Z-scores were calculated for each protein across the five timepoints (mean 0, SD 1) from cohort means that were not stratified by outcome group. **(D)** Distribution of month-3 log^2^ fold changes (cGVHD − PIT) by trajectory cluster. Negative values indicate higher abundance in PIT and positive values higher abundance in cGVHD. Violin plots show distributions with overlaid boxplots.

## Discussion

In this longitudinal proteomic study, we identified a coordinated Month-3 program that distinguishes pediatric patients who later achieve PIT from those who develop cGvHD. The divergence preceded clinical cGvHD in every affected patient (median onset 3.8 months, close to the classical day-100 threshold), so it is unlikely to reflect established disease alone. Differential-abundance, pathway and trajectory analyses pointed to a PIT-associated signature of protein degradation and folding. Across the first year, complement/coagulation, proteostasis/humoral and antigen-presentation programs predominated at different intervals. Together, these findings suggest that the plasma of the two groups already differs about three months after transplant, before clinical diagnosis.

These findings extend previous work describing impaired thymopoiesis, delayed naïve T-cell recovery, and B-cell dysregulation in cGvHD by placing these features within a longitudinal plasma-proteomic context.^19^ Rather than identifying a single dominant biomarker, the PIT-cGvHD contrast spans several molecular programs whose relative prominence changes over time. The strongest PIT-associated signal at Month 3 involved proteins related to proteasome function, protein folding and ubiquitin-dependent protein turnover. All eight subunits of the TCP1 ring complex (CCT/TRiC), which folds actin and tubulin and supports TCR-driven immune-synapse organization,^20,21^ were also higher in PIT (**Supplementary Table S2**). The proteasome regulates antigen processing, NF-κB-dependent inflammatory signaling and lymphocyte survival,^22-24^ and its pharmacological inhibition can modulate alloimmune responses. In mice, bortezomib given immediately after transplantation inhibited acute GvHD, whereas administration five or more days later accelerated GvHD-dependent morbidity,^24,25^ and it has been tested clinically as GvHD prophylaxis.^26^ These observations concern proteasome activity within cells and do not predict the direction of plasma abundance. Proteasomes are also detectable in plasma, where their abundance changes in several inflammatory conditions.^27,28^ Because these are intracellular proteins, their plasma abundance may reflect protein turnover or release from damaged or activated cells, rather than a secreted function.^27,28^

A more active, recovering hematopoietic and lymphoid compartment is one plausible explanation. It would fit the nominal association of the PIT pathway score with recent thymic emigrants and naïve CD4^+^ T cells, and it agrees with work linking thymic recovery and naïve T-cell restoration to favorable immune recovery after HSCT.^6,10^ However, mixed-effects models did not show definitive differential reconstitution, and the correlations were exploratory, based on limited paired data and not adjusted for multiple testing. We therefore hypothesize that appropriately regulated protein turnover in this early window accompanies the establishment of immune tolerance. Because we measured plasma protein abundance and not activity, cell origin or function, this and the interpretation below remain hypothesis-generating.

This pattern is consistent with a transient period of coordinated post-transplant recovery in PIT, rather than a stable proteomic state that persists throughout the first year after transplantation. On the cGvHD side, enrichment of platelet activation, cell adhesion, and transmembrane transport pathways may be consistent with vascular inflammation and endothelial dysfunction implicated in GvHD.^29^ Platelet- and stroma-derived growth factors, including PDGF and TGF-β, have been implicated in the later fibrotic phase of the disease, which makes early platelet- and adhesion-related signals biologically plausible,^30,31^ although we did not measure these factors. Together, these findings suggest that, by Month 3, tolerance may be supported by increased protein turnover and quality control, whereas the path to cGvHD is supported by active early vascular and tissue stress.^17,32^

The value of these findings lies in timing and pattern, not in any single molecule. If confirmed, this would indicate that the plasma environment diverges before clinical diagnosis, when intervention might still be considered. However, the study was not designed for prospective prediction, and validation in an independent cohort with pre-specified predictive modelling and clinically relevant performance measures is required.^33,34^ The pathways also yield testable questions: whether early platelet- and adhesion-related signaling contributes to later fibrotic remodeling, and whether proteasome- and chaperonin-related abundance marks the establishment of immune tolerance. We do not propose a biomarker or therapeutic target; Month 3 may represent an early branching point between tolerance and chronic disease that warrants further study in larger independent pediatric cohorts.

Several analyses support the internal consistency of the Month-3 signal. The PIT-associated signature remained nominally higher in PIT when re-derived without the scored patient under LOPO cross-validation, indicating that separation was not solely due to using the same samples for construction and evaluation, although the margin was narrow. The reciprocal cGvHD signature did not retain significance, which may reflect heterogeneity in organ involvement, onset timing, and severity,^35,36^ or limited power and noise in a small cohort. Month-3 fold changes differed across outcome-blinded trajectory clusters, showing that the group difference is embedded in broader temporal patterns and not attributable to a few isolated proteins. Because cluster assignment partly reflects each protein”s Month-3 shape, this is consistency and not independent replication. A linear group×time GEE model detects sustained average differences and found seven proteins rising more steeply in cGvHD (**Supplementary Table S1, Supplementary Fig. S6**). Because time was modeled linearly, it was not designed to detect a transient Month 3 signal and neither supports nor contradicts it. Month-3 was chosen after screening all timepoints, so its prominence may partly reflect selection. The same cross-validated procedure separated the groups at Month 3 but not at Month 1 or 6, which is consistent with a time-localized signal, although a difference in significance between timepoints is not a formal test of timepoint specificity.

The plasma signal should not be interpreted as a direct readout of circulating leukocyte expression alone. Several representative proteins (**Fig. 4C**), including CEMIP, CFI, and CHRD, were not detected in sorted immune-cell populations (**Supplementary Fig. S8, Supplementary Table S6**) and may originate from hepatic, stromal, endothelial, or other non-leukocyte sources. The Month-3 plasma proteomic program may therefore integrate immune-cell activity with systemic tissue responses to transplantation, including vascular, metabolic, and tissue-remodeling processes.^37^ This is particularly relevant in cGvHD, a multisystem disorder in which immune dysregulation, endothelial injury, and tissue damage may coexist.^36^ Because proteasome, chaperonin and related proteins are also abundant in red cells and platelets, contributions from blood-cell-derived material cannot be excluded.

Several limitations merit consideration. The cohort was modest, especially for timepoint-specific and paired proteomic-flow analyses, limiting power and covariate adjustment. Sampling was incomplete, so residual bias cannot be excluded. Month 3 was selected as the timepoint of strongest separation, so effect sizes may be inflated. Prior acute GvHD could not be adjusted for because it is confounded with the outcome definition. The Month-3 signature therefore distinguishes complete primary tolerance from the trajectory leading to cGvHD and is not a cGvHD-specific signature, and cohorts including secondary acute GvHD are needed. Donor type and conditioning intensity differed modestly across groups and were not covariates, so residual confounding cannot be excluded. LC-MS coverage misses low-abundance cytokines and chemokines that affinity-based assays quantify better, so non-detection of an established biomarker is not evidence against its relevance.

Despite these limitations, the study combines repeated plasma proteomics with longitudinal immune phenotyping in a pediatric allo-HSCT cohort, and integrates timepoint-specific, pathway-level, LOPO-validated, outcome-blinded trajectory and longitudinal analyses. Together, these analyses nominate Month 3 as a potentially informative interval of early molecular divergence after transplantation, and an exploratory PIT-associated plasma signature of protein degradation and folding for prospective validation.

## Data Availability

All data produced in the present study are available as follows. Processed protein-level abundance data and all differential-abundance, pathway-enrichment and trajectory-cluster results are provided in the Supplementary Tables. The raw mass spectrometry files derive from pseudonymised pediatric clinical samples and cannot be released openly because of the risk of participant re-identification. They will be deposited in a national controlled-access archive for human research data (accession number to be added), with access requests evaluated by an independent data access committee. Until then, they are available from the corresponding author on reasonable request under a data-access agreement. Individual-level clinical and immune-phenotyping data are available on reasonable request, subject to ethics approval and data-protection requirements. The Olink data were previously published (reference 18), and the Human Protein Atlas immune-cell transcriptomic data used in the supplementary analysis are publicly available.

## Acknowledgments

We thank the patients and their families for participating in this study, and the clinical and laboratory staff of the Children”s Hospital and the HUS Diagnostic Center, Helsinki University Hospital, for sample collection and processing.

## Contributions

**DK:** Conceptualization; data curation; formal analysis; investigation; project administration; visualization; writing – original draft, review, and editing. **EA** and **HL:** Clinical data collection. **YJ, EJ** and **MV** performed the proteomics. **AA, SR** and **MK:** Patient recruitment and data collection. **EK:** Conceptualization, funding acquisition, project administration, supervision. All authors reviewed the analyses, contributed to interpretation of results and writing of the manuscript, and approved the final version of the submitted report.

## Data availability statement

Processed protein-level abundance data are provided in the Supplementary Data. The raw mass spectrometry files derive from pseudonymised pediatric clinical samples and cannot be released openly because of the risk of participant re-identification. They will be deposited in a national controlled-access archive for human research data, with access requests evaluated by an independent data access committee. Until then, they are available from the corresponding author on reasonable request under a data-access agreement.

## References

1. Shah R, Murphy D, Logue M, et al. Multidisciplinary Management of Morbidities Associated with Chronic Graft-Versus-Host Disease. Clin Hematol Int. 2024;6(4):74–88. doi:10.46989/001c.124926

2. Zeiser R, Blazar BR. Pathophysiology of Chronic Graft-versus-Host Disease and Therapeutic Targets. N Engl J Med. Dec 28 2017;377(26):2565–2579. doi:10.1056/NEJMra1703472

3. Jagasia MH, Greinix HT, Arora M, et al. National Institutes of Health Consensus Development Project on Criteria for Clinical Trials in Chronic Graft-versus-Host Disease: I. The 2014 Diagnosis and Staging Working Group report. Biol Blood Marrow Transplant. Mar 2015;21(3):389–401 e1. doi:10.1016/j.bbmt.2014.12.001

4. Ng B, Harris AC, Abdossamadi S, et al. Identification of Distinct Subtypes in Immune Tolerance after Hematopoietic Cell Transplantation Using the Prospective ABLE1.0 Pediatric Study Cohort. Transplant Cell Ther. Jan 2026;32(1):84 e1–84 e16. doi:10.1016/j.jtct.2025.09.034

5. Meyer T, Maas-Bauer K, Wasch R, et al. Immunological reconstitution and infections after alloHCT - a comparison between post-transplantation cyclophosphamide, ATLG and non-ATLG based GvHD prophylaxis. Bone Marrow Transplant. Mar 2025;60(3):286–296. doi:10.1038/s41409-024-02474-1

6. Yanir A, Schulz A, Lawitschka A, Nierkens S, Eyrich M. Immune Reconstitution After Allogeneic Haematopoietic Cell Transplantation: From Observational Studies to Targeted Interventions. Front Pediatr. 2021;9:786017. doi:10.3389/fped.2021.786017

7. Jiang P, Zhou X, Cai Y, et al. Lymphocyte subset reconstitution and clinical outcomes following haploidentical hematopoietic stem cell transplantation. Br J Cancer. May 2026;134(9):1289–1299. doi:10.1038/s41416-026-03345-w

8. Gafken PR, Paczesny S. Blood proteomics for quantitative biomarkers of cellular therapies. Biomark Res. Sep 29 2025;13(1):120. doi:10.1186/s40364-025-00837-4

9. Kim DJ, Lee YJ, Choi JW, et al. Early fecal metabolomic profiling for predicting acute graft-versus-host disease following allogeneic hematopoietic stem cell transplantation. Sci Rep. Mar 6 2026;16(1)doi:10.1038/s41598-026-38818-8

10. Velardi E, Dudakov JA, van den Brink MR. Clinical strategies to enhance thymic recovery after allogeneic hematopoietic stem cell transplantation. Immunol Lett. Sep–Oct 2013;155(1-2):31–5. doi:10.1016/j.imlet.2013.09.016

11. Jarvinen E, Liu X, Varjosalo M, Keskitalo S. Automated Mag-Net Enrichment Unlocks Deep and Cost-Effective LC-MS Plasma Proteomics. J Proteome Res. Feb 6 2026;25(2):618–632. doi:10.1021/acs.jproteome.5c00420

12. Aarimaa E, Kesalainen A, Askeli S, et al. Evaluating Pediatric Reference Ranges for Extended Immunophenotyping from a Finnish Cohort against Published References. J Clin Immunol. Nov 18 2025;45(1):162. doi:10.1007/s10875-025-01959-y

13. Wu T, Hu E, Xu S, et al. clusterProfiler 4.0: A universal enrichment tool for interpreting omics data. Innovation (Camb). Aug 28 2021;2(3):100141. doi:10.1016/j.xinn.2021.100141

14. Ritchie ME, Phipson B, Wu D, et al. limma powers differential expression analyses for RNA-sequencing and microarray studies. Nucleic Acids Res. Apr 20 2015;43(7):e47. doi:10.1093/nar/gkv007

15. Halekoh U, Hojsgaard S, Yan J. The R Package geepack for Generalized Estimating Equations. J Stat Softw. Jan 2006;15(2):1–11. doi:DOI 10.18637/jss.v015.i02

16. Wu X, Qian J, Tong H, Huang X. Bridging Innate and Adaptive Immunity: Understanding and Targeting the Complement System in GVHD Pathogenesis and Therapy. Transplant Cell Ther. Feb 2026;32(2):141–155. doi:10.1016/j.jtct.2025.10.014

17. Neidemire-Colley L, Robert J, Ackaoui A, Dorrance AM, Guimond M, Ranganathan P. Role of endothelial cells in graft-versus-host disease. Front Immunol. 2022;13:1033490. doi:10.3389/fimmu.2022.1033490

18. Alexandersson A, Venalainen MS, Heikkila N, et al. Proteomics screening after pediatric allogenic hematopoietic stem cell transplantation reveals an association between increased expression of inhibitory receptor FCRL6 on gammadelta T cells and cytomegalovirus reactivation. Immunol Cell Biol. Jul 2024;102(6):513–525. doi:10.1111/imcb.12762

19. Dekker L, Sanders E, Lindemans CA, de Koning C, Nierkens S. Naive T Cells in Graft Versus Host Disease and Graft Versus Leukemia: Innocent or Guilty? Front Immunol. 2022;13:893545. doi:10.3389/fimmu.2022.893545

20. Martin-Cofreces NB, Chichon FJ, Calvo E, et al. The chaperonin CCT controls T cell receptor-driven 3D configuration of centrioles. Sci Adv. Dec 2020;6(49)doi:10.1126/sciadv.abb7242

21. Wu C, Jiang M, Yang X, et al. ANKRD55 is a key regulator of T cell inflammation in multiple sclerosis. J Clin Invest. Oct 15 2025;135(20)doi:10.1172/JCI195214

22. Kloetzel PM. The proteasome and MHC class I antigen processing. Biochim Biophys Acta. Nov 29 2004;1695(1-3):225–33. doi:10.1016/j.bbamcr.2004.10.004

23. Wang Y, Liang Y, Zhang Y, Wu D, Liu H. Bortezomib inhibits bone marrow-derived dendritic cells. Int J Clin Exp Pathol. 2015;8(5):4857–62.

24. Sun K, Welniak LA, Panoskaltsis-Mortari A, et al. Inhibition of acute graft-versus-host disease with retention of graft-versus-tumor effects by the proteasome inhibitor bortezomib. Proc Natl Acad Sci U S A. May 25 2004;101(21):8120–5. doi:10.1073/pnas.0401563101

25. Sun K, Wilkins DE, Anver MR, et al. Differential effects of proteasome inhibition by bortezomib on murine acute graft-versus-host disease (GVHD): delayed administration of bortezomib results in increased GVHD-dependent gastrointestinal toxicity. Blood. Nov 1 2005;106(9):3293–9. doi:10.1182/blood-2004-11-4526

26. Koreth J, Stevenson KE, Kim HT, et al. Bortezomib, tacrolimus, and methotrexate for prophylaxis of graft-versus-host disease after reduced-intensity conditioning allogeneic stem cell transplantation from HLA-mismatched unrelated donors. Blood. Oct 29 2009;114(18):3956–9. doi:10.1182/blood-2009-07-231092

27. Zoeger A, Blau M, Egerer K, Feist E, Dahlmann B. Circulating proteasomes are functional and have a subtype pattern distinct from 20S proteasomes in major blood cells. Clin Chem. Nov 2006;52(11):2079–86. doi:10.1373/clinchem.2006.072496

28. Egerer K, Kuckelkorn U, Rudolph PE, et al. Circulating proteasomes are markers of cell damage and immunologic activity in autoimmune diseases. J Rheumatol. Oct 2002;29(10):2045–52.

29. Luft T, Dreger P, Radujkovic A. Endothelial cell dysfunction: a key determinant for the outcome of allogeneic stem cell transplantation. Bone Marrow Transplant. Oct 2021;56(10):2326–2335. doi:10.1038/s41409-021-01390-y

30. Olivieri A, Locatelli F, Zecca M, et al. Imatinib for refractory chronic graft-versus-host disease with fibrotic features. Blood. Jul 16 2009;114(3):709–18. doi:10.1182/blood-2009-02-204156

31. Olivieri A, Cimminiello M, Corradini P, et al. Long-term outcome and prospective validation of NIH response criteria in 39 patients receiving imatinib for steroid-refractory chronic GVHD. Blood. Dec 12 2013;122(25):4111–8. doi:10.1182/blood-2013-05-494278

32. Lelas A, Greinix HT, Wolff D, Eissner G, Pavletic SZ, Pulanic D. Von Willebrand Factor, Factor VIII, and Other Acute Phase Reactants as Biomarkers of Inflammation and Endothelial Dysfunction in Chronic Graft-Versus-Host Disease. Front Immunol. 2021;12:676756. doi:10.3389/fimmu.2021.676756

33. Yu J, Storer BE, Kushekhar K, et al. Biomarker Panel for Chronic Graft-Versus-Host Disease. J Clin Oncol. Aug 1 2016;34(22):2583–U57. doi:10.1200/Jco.2015.65.9615

34. Paczesny S, Hakim FT, Pidala J, et al. National Institutes of Health Consensus Development Project on Criteria for Clinical Trials in Chronic Graft-versus-Host Disease: III. The 2014 Biomarker Working Group Report. Biol Blood Marrow Tr. May 2015;21(5):780–792. doi:10.1016/j.bbmt.2015.01.003

35. Hill GR, Betts BC, Tkachev V, Kean LS, Blazar BR. Current Concepts and Advances in Graft-Versus-Host Disease Immunology. Annu Rev Immunol. Apr 26 2021;39:19– 49. doi:10.1146/annurev-immunol-102119-073227

36. Cooke KR, Luznik L, Sarantopoulos S, et al. The Biology of Chronic Graft-versus-Host Disease: A Task Force Report from the National Institutes of Health Consensus Development Project on Criteria for Clinical Trials in Chronic Graft-versus-Host Disease. Biol Blood Marrow Transplant. Feb 2017;23(2):211–234. doi:10.1016/j.bbmt.2016.09.023

37. Malmstrom E, Malmstrom L, Hauri S, et al. Human proteome distribution atlas for tissue-specific plasma proteome dynamics. Cell. May 15 2025;188(10):2810–2822 e16. doi:10.1016/j.cell.2025.03.013

