## Supplementary Figures for "Longitudinal Plasma Proteomics Distinguishes Primary Immune Tolerance from Chronic Graft-versus-Host Disease After Pediatric allo-HSCT"

### Supplementary Figure 1

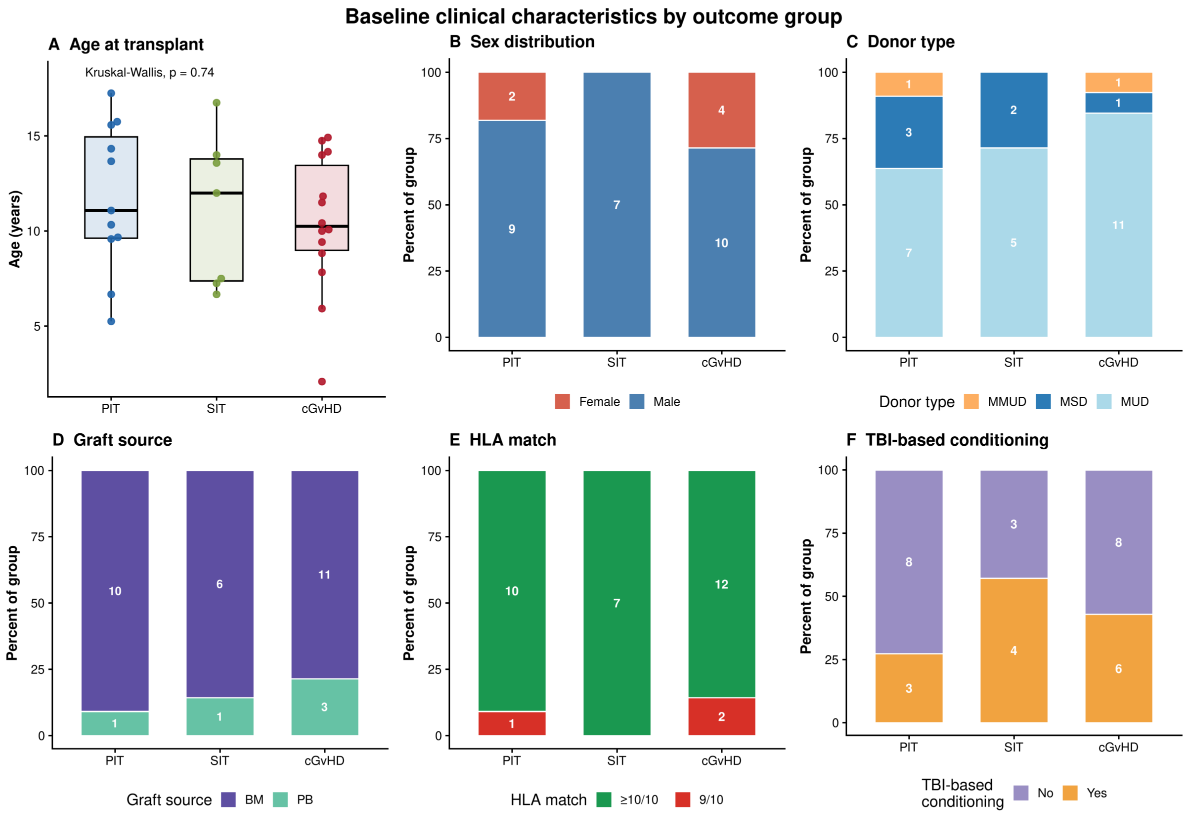


**Supplementary Figure S1. Baseline clinical characteristics by outcome group.** Transplant-related characteristics of patients with post-transplant immune tolerance (PIT, n = 11), stable immune tolerance (SIT, n = 7) and chronic graft-versus-host disease (cGvHD, n = 14). (**A**) Age at transplant (years); points show individual patients, boxes show the median and interquartile range, and whiskers extend to 1.5 × IQR. Groups were compared with the Kruskal–Wallis test (p = 0.74). (**B**) Sex distribution. (**C**) Donor type: matched unrelated donor (MUD), matched sibling donor (MSD) or mismatched unrelated donor (MMUD). (**D**) Graft source: bone marrow (BM) or peripheral blood (PB). (**E**) HLA match: ≥10/10 or 9/10. (**F**) Total body irradiation (TBI)-based conditioning (yes or no). In panels B–F, bars show the percentage of patients within each group and the numbers inside the bars show patient counts.

### Supplementary Figure S2

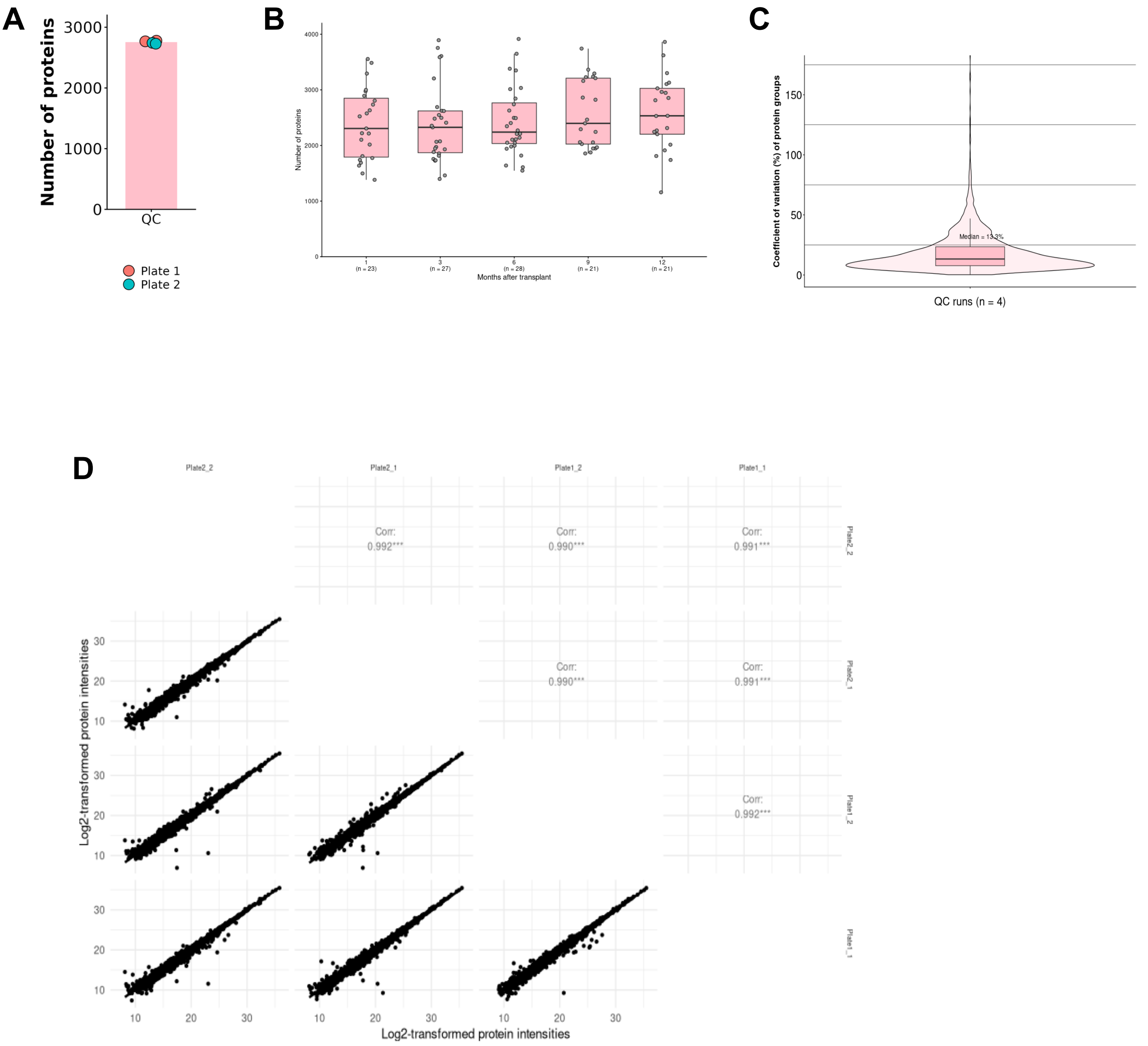


**Supplementary Figure S2. Proteomics quality control and identification depth. (A)** Number of protein groups identified in each of four QC runs (two plates, two replicate injections per plate); bars show the mean and points show individual runs, coloured by plate. **(B)** Number of proteins identified per cohort sample at 1–12 months after transplant (n per timepoint shown below the axis); boxes show the median and interquartile range, and points are individual samples. **(C)** Coefficient of variation (CV) of protein-group intensities across the QC runs, shown as a violin plot with an overlaid boxplot. **(D)** Pairwise correlation (Pearson) of log2-transformed protein intensities between QC runs. In the QC runs a mean of 2,755 protein groups was identified (range 2,729–2,778), with a median CV of 13.3% (below 15%), and QC runs correlated well between and within plates, indicating low technical variability. Across the cohort, 4,702 proteins were quantified, with a median of 2,336 identifications per sample (range 1,157–3,915; 120 samples).

### Supplementary Figure S3

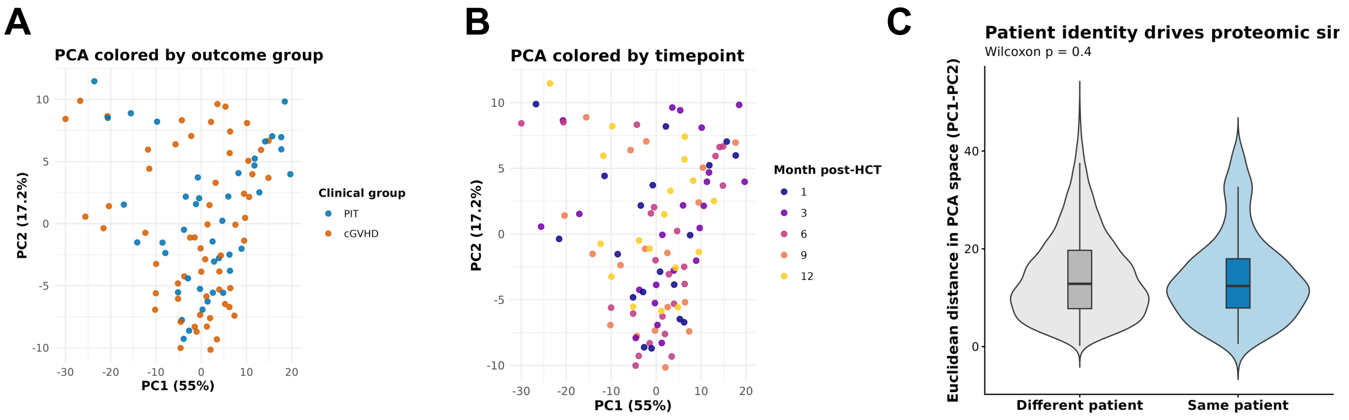


**Supplementary Figure S3**. **(A) Global PCA colored by clinical outcome.** PCA of the top 200 most variable plasma proteins (log2-transformed intensities), including all longitudinal samples from patients with primary immune tolerance (PIT, blue) or chronic graft-versus-host disease (cGVHD, orange). Each point represents one patient-timepoint sample. PIT and cGVHD samples overlap substantially across the projection, indicating that clinical outcome is not the dominant axis of proteomic variance. **(B) Global PCA colored by post-transplant timepoint.** Same PCA projection as panel A, with samples colored by month post-HCT (1, 3, 6, 9, or 12). Samples do not separate cleanly by timepoint either, indicating that follow-up time alone does not organize the dominant PCA axes. **(C) Within- versus between-patient distances.** Pairwise Euclidean distances in PC1–PC2 space between samples from the same patient (n = 170 pairs) and from different patients (n = 4,780 pairs), in the PCA of the 200 most variable proteins. Same-patient pairs were not significantly closer than different-patient pairs (median 12.5 vs. 12.9; Wilcoxon rank-sum two-sided P = 0.41; pairs are not independent, so the P-value is approximate). Violins show the distribution, and boxes show the median and interquartile range.

### Supplementary Figure S4

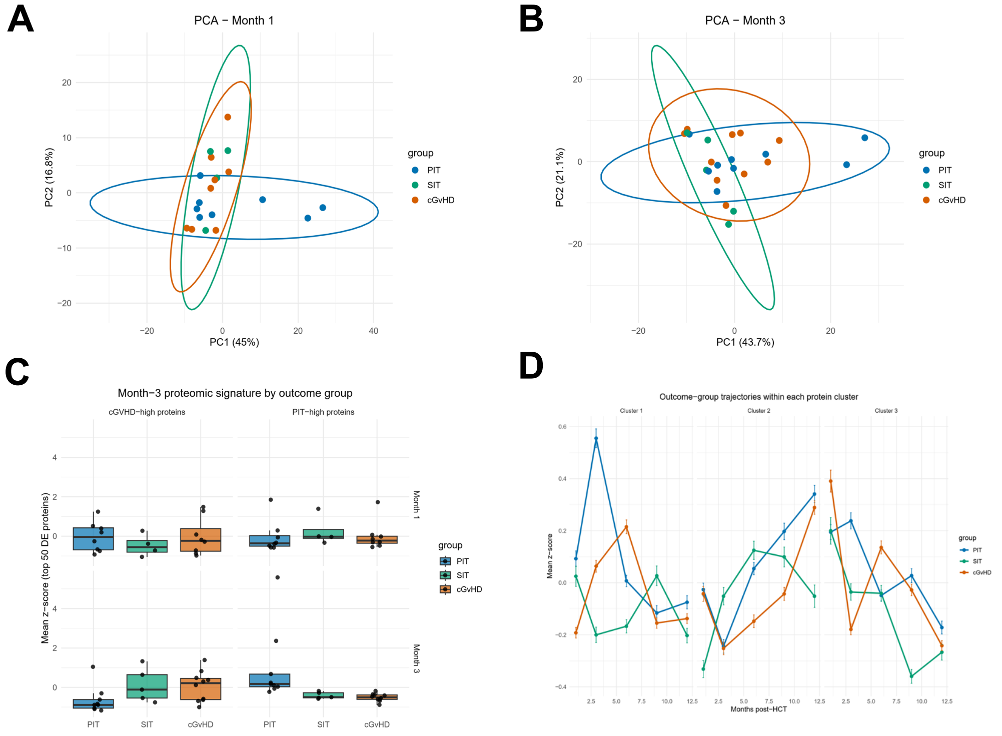


**Supplementary Figure S4. Exploratory proteomic characterization of PIT, SIT, and cGVHD outcomes.** **(A,B)** Principal component analysis of $log_2$-transformed plasma-protein abundances at Months 1 and 3 post-transplant. Each point represents one patient sample; ellipses indicate 95% confidence regions. Percentages of variance explained by PC1 and PC2 are shown in parentheses. **(C)** Exploratory proteomic signature scores at Months 1 and 3. For each direction, scores were calculated as the mean protein-level z-score across the 50 proteins with the smallest nominal P values in the Month-3 PIT-versus-cGvHD differential-abundance analysis, separately for proteins higher in PIT and proteins higher in cGvHD. Boxes show the median and interquartile range; whiskers extend to 1.5× the interquartile range; points represent individual patient samples. **(D)** Mean standardized protein-abundance trajectories within the three outcome-blinded k-means trajectory clusters (k=3), stratified by clinical outcome. Lines and points show group means ± standard error. Z-scores were calculated separately for each protein across all available samples. Analyses including SIT were descriptive; no formal SIT-specific differential-abundance testing was undertaken because of the limited SIT sample size. These signatures were derived from the same Month-3 comparison and are displayed to contextualize SIT descriptively, not as independent validation.

### Supplementary Figure S5

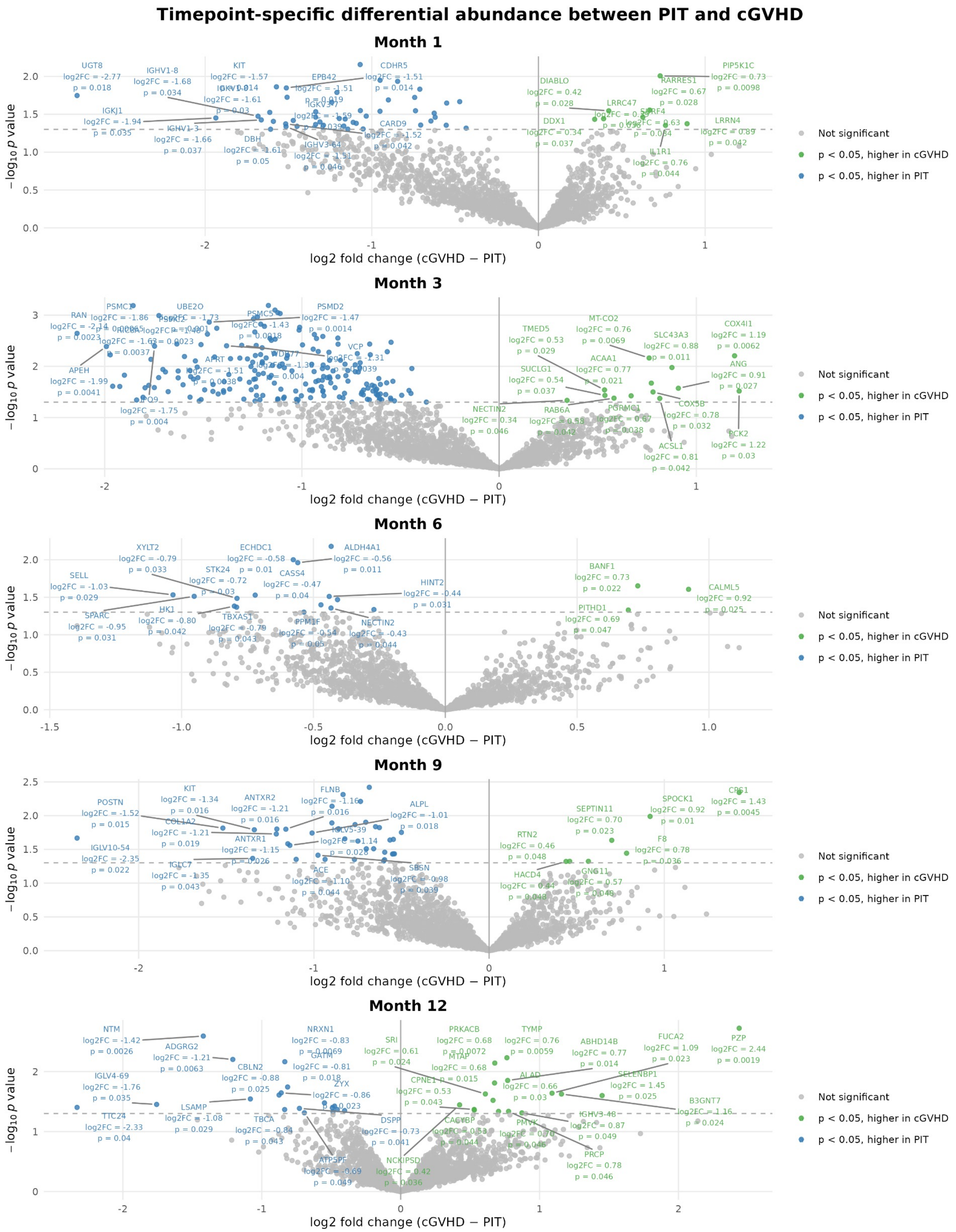


**Supplementary Figure S5. Timepoint-specific differential abundance between PIT and cGVHD at all timepoints.** Volcano plots show $\log_2$ fold change (cGVHD − PIT) vs $-\log_{10}$ P value for months 1, 3, 6, 9, and 12. Proteins detected in ≥70% of samples at each timepoint were tested using limma with empirical-Bayes moderation; remaining missing values were imputed using the protein-specific median.

### Supplementary Figure S6

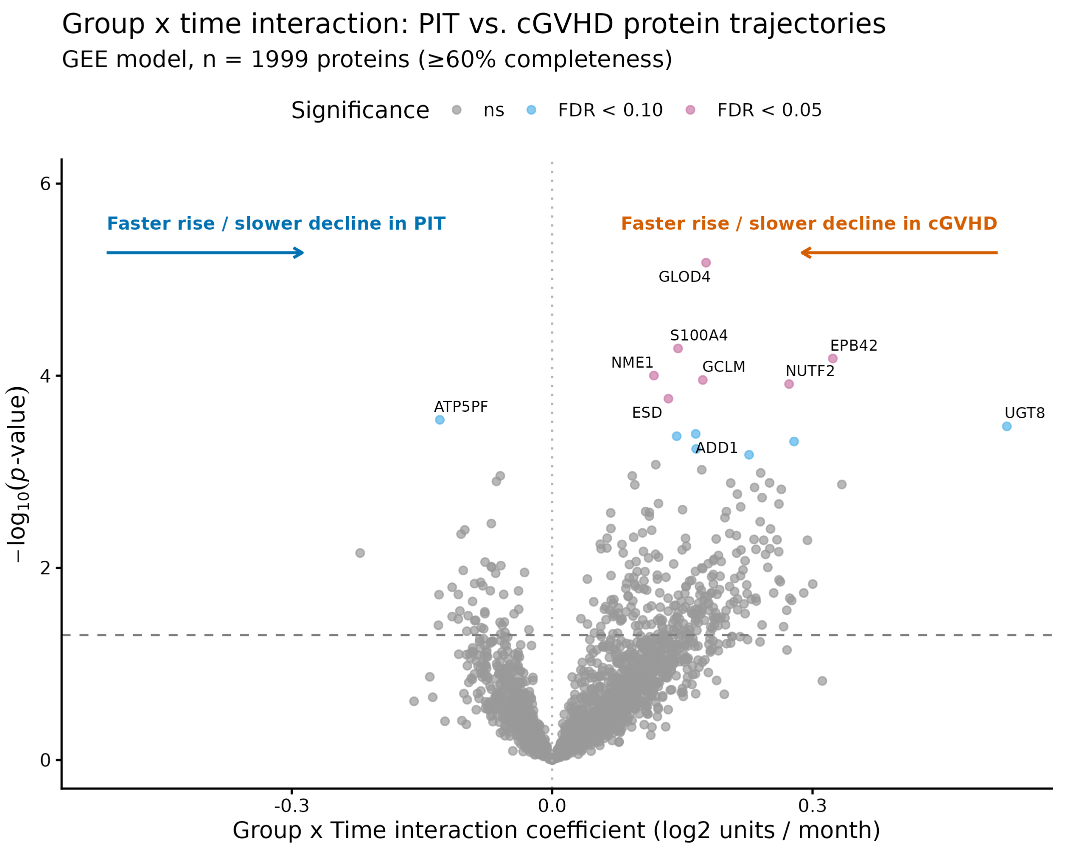


**Supplementary Figure S6. Group×time interaction analysis of protein trajectories across follow-up.** Volcano plot of generalized estimating equation (GEE) models testing a group×time interaction for each protein with sufficient longitudinal completeness (≥60% of possible measurements across Months 1, 3, 6, 9, and 12; n = 1,999 of 4,702 quantified proteins; Methods). PIT is the reference group. The x-axis shows the interaction coefficient (log2 units/month); positive values indicate a more positive slope in cGvHD than in PIT (steeper rise or slower decline in cGvHD), and negative values indicate the reverse. The y-axis shows $-\log_{10}$(p-value) from the Wald test on the interaction term. Points are colored by Benjamini–Hochberg FDR-adjusted significance (FDR < 0.05, FDR < 0.10, or not significant); the dashed horizontal line marks p = 0.05, and the dotted vertical line marks a null interaction effect. The ten most significant proteins are labeled. Seven proteins reached FDR < 0.05, all with a more positive slope in cGvHD than in PIT. Because this test assumes a linear trend over time, it is complementary to, rather than a replacement for, the transient Month-3 signal identified by timepoint-specific differential expression and outcome-blinded trajectory clustering (**Figs. 1C, 2, 4**).

### Supplementary Figure S7

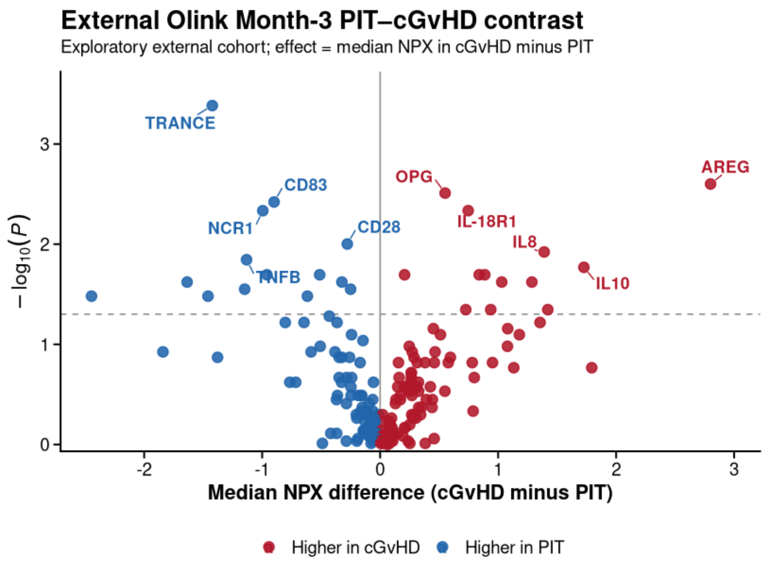


**Supplementary Figure S7. Exploratory Month-3 PIT–cGvHD protein differences in an external Olink cohort.** Volcano plot of the Month-3 comparison between patients classified as primary immune tolerance (PIT, n = 19) and chronic graft-versus-host disease (cGvHD, n = 6) in an external pediatric allo-HSCT cohort profiled with Olink Inflammation and Immune Response panels (180 proteins). The x-axis shows the difference in median normalized protein expression (NPX) between cGvHD and PIT; positive values indicate higher protein abundance in cGvHD and negative values indicate higher abundance in PIT. The y-axis indicates $-\log_{10}$ nominal two-sided Wilcoxon rank-sum test P values. Points are colored by direction of the group contrast, and the ten proteins with the smallest nominal P values are labeled. The dashed line denotes nominal P=0.05. Twenty-seven proteins had nominal P<0.05; none reached a Benjamini–Hochberg-adjusted P<0.05 (smallest adjusted P=0.074). Results are exploratory and were not used to define the LC–MS discovery signature.

### Supplementary Figure S8

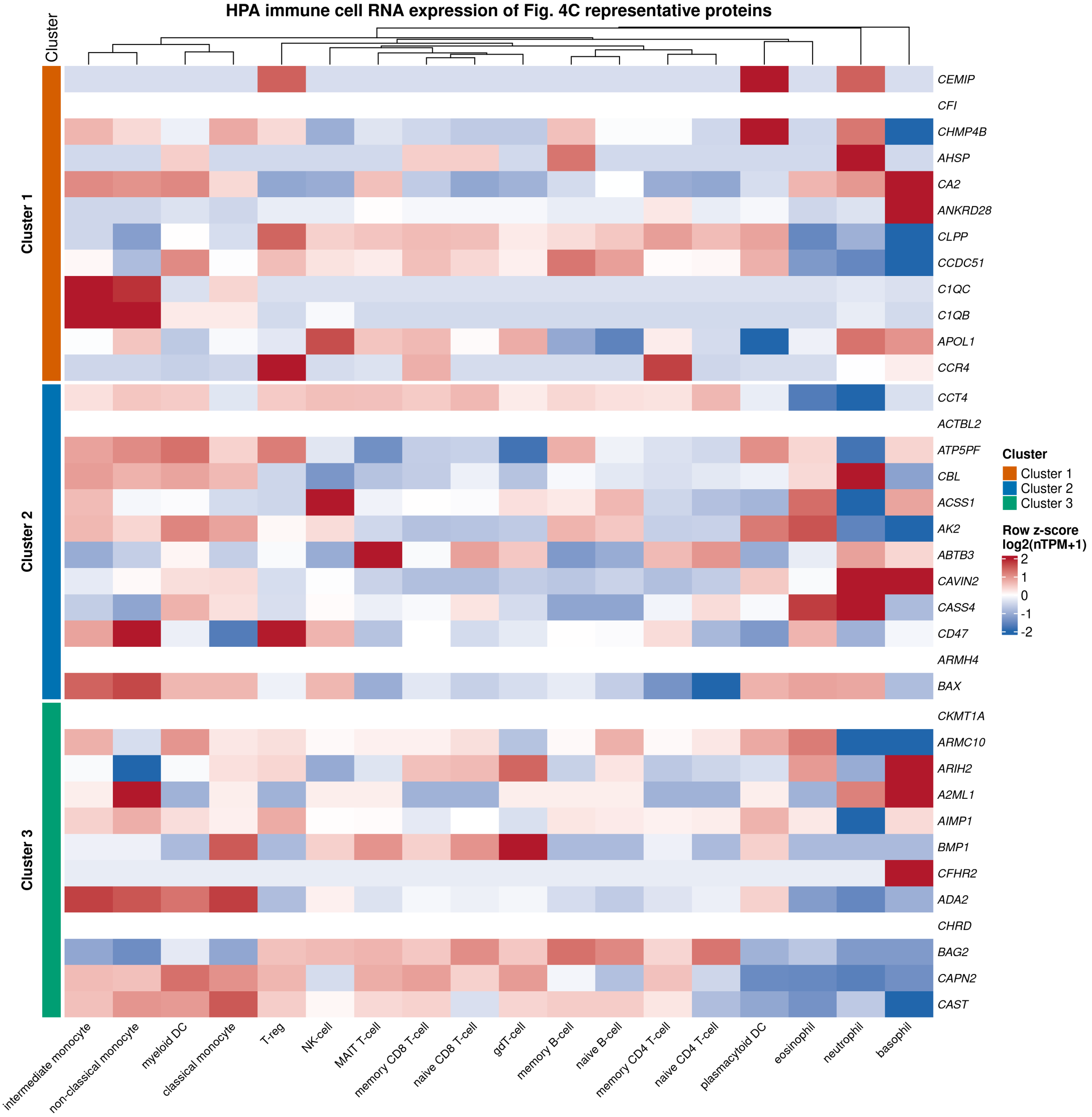


**Supplementary Figure S8. Immune cell RNA expression of Fig. 4C representative proteins.** Heatmap of RNA expression (Human Protein Atlas, 18 sorted immune cell types) for the 36 proteins shown in Fig. 4C (top 12 per trajectory cluster, ranked by trajectory shape score). Expression values are log2(nTPM+1)-transformed and row-scaled (z-score) across cell types to highlight relative cell-type specificity per protein. Rows are grouped by trajectory cluster (Cluster 1, orange; Cluster 2, blue; Cluster 3, green; left annotation bar) and columns are hierarchically clustered by expression similarity across proteins. Red indicates relatively higher expression, blue indicates relatively lower expression, within each protein's own row. Fisher's exact tests comparing the proportion of proteins assigned to each immune cell lineage (B-cell, T-cell, NK-cell, monocyte, granulocyte, dendritic cell, or not detected) within each cluster against the full annotated Human Protein Atlas gene background did not reach statistical significance for any cluster–lineage pair (all $P > 0.05$; **Supplementary Table S6**), although Cluster 3 showed a nominal enrichment for monocyte-lineage expression (25% vs. 9% background; odds ratio 3.4; $P = 0.09$, uncorrected). Several proteins (e.g., CEMIP, CFI, CHRD, CKMT1A, A2ML1, BMP1, CFHR2, ARMH4, ACTBL2) were not detected in any sorted circulating immune cell population, consistent with non-leukocyte (e.g., hepatic, stromal, or platelet-derived) origins.
