## Supplementary Methods for "Longitudinal Plasma Proteomics Distinguishes Primary Immune Tolerance from Chronic Graft-versus-Host Disease After Pediatric allo-HSCT"

### Plasma sample collection and LC-MS proteomics

Peripheral blood plasma samples were collected at each timepoint in EDTA-coated tubes. Peripheral blood mononuclear cell isolation was performed by gradient centrifugation (Ficoll-Paque PLUS, Cytiva, Buckinghamshire, UK). Plasma was separated by centrifugation at 2000 × g for 15 min and stored at −80°C until analysis. Patients were scheduled for longitudinal sampling at Months 1, 3, 6, 9, and 12 post-transplant; actual sampling completeness varied, with 14 of 32 patients sampled at all five timepoints and 18 patients sampled at 1–4 timepoints (median, 4 timepoints per patient). Month-1 samples were most frequently unavailable. In total, 120 samples were collected and analyzed.

Plasma proteins from 20 μL of plasma were enriched using an automated Mag-Net plasma enrichment workflow, and the resulting peptides were analyzed by liquid chromatography-mass spectrometry (LC-MS). The fully automated Mag-Net workflow and the LC-MS untargeted proteomics method with Evosep One (Odense, Denmark), employing the 60-samples-per-day method, connected to Orbitrap Astral (Thermo Fisher Scientific, Waltham, MA, USA), have been previously reported in detail (*Jarvinen et al. J Proteome Res 25, 2026 Feb 6;25(2):618-632*)

Method robustness was ensured by processing 2 quality control (QC) samples per plate (**Supplementary Figure S2**), consisting of pooled commercial reference plasma (Research Donors Ltd, London, UK) processed alongside patient samples on the same plates. Across four QC runs (two plates, two replicate injections per plate), a mean of 2,754 protein groups was identified (range, 2,729–2,778), with a median coefficient of variation of 13.3% across 2,909 protein groups, confirming low plate-to-plate technical variability. In total, 4,702 proteins were quantified across the patient cohort, with a median of 2,336 protein identifications per sample (range, 1,157–3,915) across 120 patient samples.

### Flow cytometry and immune cell count

Sample preparation followed a previously described procedure (*Alexandersson et al. Immunol Cell Biol. 2024 Jul;102(6):513-525*). Briefly, frozen peripheral blood mononuclear cells (PBMCs) were thawed in RPMI-based thawing solution, washed, and resuspended in staining buffer. Cells were incubated with a fluorochrome-conjugated antibody cocktail and LIVE/DEAD Fixable Green Dead Cell Stain (Thermo Fisher Scientific) in Brilliant Stain Buffer (BD Biosciences) for 30 min at 4°C.

Data were acquired on an LSR II Fortessa flow cytometer (BD Biosciences) and analyzed using FlowJo software (v10.10.1; BD Biosciences).

Peripheral blood immune cell phenotyping was performed at each timepoint using an extended immunophenotyping panel encompassing T-cell markers (CD3, CD4, CD8, CD45RA, CCR7, CD31, TCRαβ, TCRγδ), regulatory and activation markers (FoxP3-defined Treg, HLA-DR, CD38), B-cell markers (CD19, naive, memory, non-switched memory, switched memory, activated, transitional, and plasmablast subsets), and NK cells ($CD56^{+}CD3^{-}$). Absolute cell counts were calculated using a dual-platform approach, combining the percentage of each immunophenotypic subset determined by flow cytometry with the absolute lymphocyte count from a concurrent automated complete blood count. Key populations included total T cells ($CD3^{+}$), $CD4^{+}$ and $CD8^{+}$ T cells, naive $CD4^{+}$ T cells ($CD45RA^{+}CCR7^{+}$), recent thymic emigrants (RTE; $CD45RA^{+}CD31^{+}CD4^{+}$), γδ T cells, and $CD19^{+}$ B cells, including naive B cells.

#### Trajectory clustering and representative proteins per cluster

For each protein, a cohort mean log2-transformed intensity was calculated at each of the five post-transplant timepoints (Months 1, 3, 6, 9, and 12), averaging across all patients with an available measurement at that timepoint, irrespective of outcome group. Proteins were retained only if a non-missing cohort-mean value was obtained at all five timepoints; no imputation was required, as timepoint-level averaging inherently accommodated per-patient missingness.

From the retained proteins, each protein's five-timepoint cohort-mean trajectory was first z-score standardized (mean 0, SD 1 across the five months), and 500 proteins were selected for downstream clustering based on this standardized representation. Because standardization equalizes variance across timepoints for any protein with non-constant abundance, this selection step functioned primarily to exclude proteins with flat or near-constant trajectories rather than to rank proteins by the magnitude of their temporal dynamics.

The 500 selected proteins were subjected to k-means clustering (k=3, nstart=100) without using outcome group labels. Cluster-level mean trajectory was computed as the mean z-score per cluster per month. To illustrate the proteins underlying each trajectory pattern, a cluster-specific shape score was computed for each protein, quantifying conformity to the cluster-typical profile: for the Month-3-peak cluster, the Month-3 z-score minus the mean z-score of the remaining timepoints; for the Month-3-nadir cluster, the mean of Months 6–12 minus the Month-3 z-score; for the early-high/late-decline cluster, the Month-1 minus Month-12 z-score. The top 12 proteins per cluster by shape score were visualized as a z-score heatmap.

**Differential expression and enrichment analyses (extended)**

Differential abundance between PIT and cGVHD was assessed independently at each timepoint using limma (*Ritchie et al. Nucleic Acids Res. 2015 Apr 20;43(7):e47*), with empirical-Bayes moderated t-statistics from an unpaired two-group model (PIT vs. cGVHD); topTable-derived p-values are two-sided. Benjamini–Hochberg FDR correction was applied within each timepoint, and proteins with FDR < 0.05 were considered significant. Proteins were tested if detected in ≥70% of samples at that timepoint; this yielded 1,545, 1,629, 1,752, 1,844, and 1,929 proteins tested at Months 1, 3, 6, 9, and 12, respectively (at Month 3, n_PIT=9, n_cGVHD=13 with available paired data).

To test whether protein trajectories diverged by outcome group across the full follow-up period rather than at isolated timepoints, generalized estimating equation (GEE) models were additionally fit (exchangeable correlation structure, patient-clustered robust standard errors) with a group×time interaction term for each protein with ≥60% completeness across the five timepoints (1,999 of 4,702 proteins). Unlike the per-timepoint limma analyses above, which required ≥70% detection within a single timepoint, this criterion required ≥60% completeness pooled across all five timepoints combined. Benjamini–Hochberg FDR correction was applied across all 1,999 tested proteins.

Prior acute GvHD status was not included as a covariate in timepoint-specific differential-abundance models, because it is definitionally confounded with outcome group (PIT patients have, by definition, no acute GvHD; 71% of cGVHD patients had prior acute GvHD), leaving no within-group variance to adjust against.

Month 3 was identified as the timepoint of strongest PIT–cGVHD separation from this initial screen across all five sampled timepoints (Fig. 2A); the deepest characterization of this signal (module-based summary scores, leave-one-patient-out cross-validation, and trajectory-cluster association) was then conducted specifically at this data-selected timepoint, while pathway enrichment was additionally examined at Month 1 and 6 for temporal context. Because Month 3 was selected as the maximum among five tested timepoints, its effect sizes may be modestly inflated relative to an a priori single-timepoint test, and the GEE group×time model above, which does not involve timepoint selection, is reported as a partially independent check on the direction, though not magnitude, of this signal.

Gene Ontology Biological Process over-representation analysis (ORA) and gene set enrichment analysis (GSEA) were performed using clusterProfiler (v.4.12.6).[14] For ORA, the background gene universe was restricted to the set of proteins tested in the corresponding timepoint-specific differential-abundance analysis (rather than the default whole-genome background), ensuring that enrichment was assessed relative to the proteins actually detectable on this platform. For GSEA, the full ranked list of tested proteins at each timepoint (ranked by moderated t-statistic) was used, such that the same timepoint-specific protein universe was implicitly represented.

**Leave-one-patient-out (LOPO) cross-validation**

For each patient, a composite "PIT pathway score" was calculated from the standardized abundance of proteins mapping to the top PIT-enriched GO biological-process pathways at the timepoint of interest. To address the circularity of using the same samples to both build and evaluate this score, leave-one-patient-out (LOPO) cross-validation was performed: the signature was re-derived excluding each patient in turn, and the held-out patient was scored independently using the per-protein mean and standard deviation from the remaining patients only.

This procedure was applied at Month 3 using both the PIT-associated and reciprocal cGvHD-associated signatures (matching the composite scores shown in **Fig. 3E** and **Fig. 4B**), and at Months 1 and 6 using the PIT-associated signatures only (matching Fig. 2C); no reciprocal cGvHD-associated signature was defined for these two timepoints in the original analysis. Group differences in LOPO-derived scores were assessed by two-sided Wilcoxon rank-sum test. Full per-patient scores and test statistics for all three timepoints are provided in **Supplementary Table S4**.

**Statistical and bioinformatic analyses**

All analyses were performed in R (v.4.6.1) using the tidyverse (v.2.0.0) for data manipulation, limma (v.3.23) for differential-abundance testing, clusterProfiler (v.4.12.6) for pathway enrichment, geepack (v.1.3.13) for generalized estimating equation models, and lme4 (v.2.0-6) for linear mixed-effects models. Figures were generated with ggplot2 (v.4.0.3) and ggrepel (v.0.9.8). Unless otherwise specified in the relevant Methods subsection, significance was assessed using Benjamini-Hochberg false discovery rate (FDR) correction, with a threshold of FDR < 0.05 for confirmatory analyses and FDR < 0.10 explicitly flagged as exploratory where used (e.g., Month-3 differential abundance). Two-sided tests were used throughout unless otherwise stated.
